# Complex Autoantibody Responses Occur Following Moderate to Severe Traumatic Brain Injury

**DOI:** 10.1101/2020.07.24.20161786

**Authors:** Edward J Needham, Oda Stoevesandt, Eric P Thelin, Henrik Zetterberg, Elisa R Zanier, Faiez Al Nimer, Nicholas J Ashton, Joanne G Outtrim, Virginia FJ Newcombe, Hani S Mousa, Joel Simren, Kaj Blennow, Zhihui Yang Z, Peter J Hutchinson, Fredrik Piehl, Adel E Helmy, Mike J Taussig, Kevin KW Wang, Joanne L Jones, David K Menon, Alasdair J Coles

**Affiliations:** Department of Clinical Neurosciences, University of Cambridge, UK; Division of Anaesthesia, Department of Medicine, University of Cambridge, UK; Cambridge Protein Arrays Ltd, Babraham Research Campus, Cambridge, UK; Department of Clinical Neuroscience, Karolinska Institutet, Stockholm, Sweden; Department of Neurovascular Diseases, Karolinska University Hospital, Stockholm, Sweden; Department of Academic Neurosurgery, Department of Clinical Neurosciences, University of Cambridge, Cambridge, UK; Department of Psychiatry and Neurochemistry, Institute of Neuroscience & Physiology, the Sahlgrenska Academy at the University of Gothenburg, Mölndal, Sweden; Clinical Neurochemistry Laboratory, Sahlgrenska University Hospital, Mölndal, Sweden; Department of Neurodegenerative Disease, UCL Institute of Neurology, Queen Square, London, UK; UK Dementia Research Institute at UCL, London, UK; Department of Neuroscience, Istituto di Ricerche Farmacologiche Mario Negri IRCCS, Milan, Italy; Wolfson Brain Imaging Centre, University of Cambridge, UK; Program for Neurotrauma, Neuroproteomics and Biomarker Research, Departments of Emergency Medicine, Psychiatry and Neuroscience, University of Florida, McKnight Brain Institute, USA

**Keywords:** Traumatic brain injury, autoantibodies, secondary injury, neurodegeneration, neuroinflammation

## Abstract

The majority of variation in outcome following severe traumatic brain injury (TBI) remains unexplained by currently recognised prognostic factors, suggesting a contribution from unaccounted variables. One key candidate variable is neuroinflammation, including the generation of autoantibodies against brain specific antigens which have been described in some individuals following TBI. Here we hypothesised that autoantibody responses following TBI would differ between individuals, and would explain a proportion of outcome variance.

We developed a custom protein microarray to characterise the generation of autoantibodies to both central nervous system and systemic antigens in the acute-phase of TBI (within ten days of injury), and to determine their late (6-12 months) and long-term (6-13 years) persistence.

We identified two distinct patterns of response. The first was a broad response to the majority of antigens tested, predominantly IgM-mediated in the acute-phase, then IgG-dominant at late and long-term time-points. The second was of dominant responses to certain antigens, most frequently myelin-associated glycopeptide (MAG), which persisted for several months post-TBI but then subsequently resolved.

Exploratory analyses suggested that patients with a greater acute IgM response experienced worse outcomes than predicted. Furthermore, late persistence of anti-MAG IgM autoantibodies correlated with serum neurofilament light concentrations, suggesting an association with ongoing neurodegeneration over the first year post-injury.

Our results show that autoantibody production occurs in some individuals following TBI, can persist for many years, and may affect patient outcome. The complexity of responses mean that conventional approaches based on measuring responses to single antigenic targets may be misleading.

## Introduction

Traumatic brain injury (TBI) is the leading cause of death and disability in young adults in the developed world (Jennett, 1996). Despite significant advances in knowledge there are few reliable early prognostic markers, and there is marked heterogeneity in outcome between individuals with seemingly similar initial primary injuries. Indeed, the best established prognostic models in TBI (such as the CRASH and IMPACT models), explain less than 40% of outcome variance (Lingsma *et al*., 2010). Secondary injury, the process where (amongst others) metabolic and inflammatory consequences of TBI cause additional neurological injury, is likely to contribute significantly to this heterogeneity, and is potentially therapeutically modifiable (Werner and Engelhard, 2007). Furthermore, TBI is now increasingly believed to trigger a chronic neurodegenerative process in a subset of patients (Ruff *et al*., 1991; Millis *et al*., 2001; Sander *et al*., 2001; Hammond *et al*., 2004; Himanen *et al*., 2006; Whitnall, 2006; Greenberg *et al*., 2008; Till *et al*., 2008; Kumar *et al*., 2009; Newcombe *et al*., 2016).

The mechanisms underlying early secondary injury and late neurodegeneration are incompletely understood, but neuroinflammation has been implicated in both processes, and represents a potential therapeutic target. Most work to date has focussed on innate immunity and microglial activation, which may persist decades after injury. The intensity of the late microglial response, in particular, appears to correlate with late functional outcome and white matter damage (Ramlackhansingh *et al*., 2011; Johnson *et al*., 2013; Scott *et al*., 2015). However to-date, therapeutic modulation of these systems has not had any clinical impact (Maas *et al*., 2006; Scott *et al*., 2018).

In addition to local innate immune activation, TBI disrupts both brain tissue and the blood-brain barrier (BBB), releasing brain antigens into the systemic circulation, and into the cervical lymph nodes via glymphatic and meningeal lymphatic systems (Plog *et al*., 2015; Absinta *et al*., 2017), generating both humoral and cellular adaptive autoimmune responses, which may be detrimental (Cox *et al*., 2006; Marchi *et al*., 2013; Zhang *et al*., 2014). The phenomenon of destructive autoimmunity triggered by central nervous system (CNS) injury is well established, with notable examples including sympathetic ophthalmia, and NMDAr encephalitis following herpes simplex encephalitis (Jr *et al*., 2017; Gelfand, 2018). As far back as the 1960s, studies of small patient cohorts have described autoantibodies to individual brain antigens following TBI (Shamreĭ, 1969; Škoda *et al*., 2006; Sorokina *et al*., 2009; Ngankam *et al*., 2011; Marchi *et al*., 2013; Zhang *et al*., 2014). Given the plethora of different brain antigens released after TBI, it is unlikely that measuring a single autoantibody captures the true extent of autoimmune response. Indeed, although the cognate antigens have been poorly characterised, past studies show that autoantibodies to multiple CNS targets are likely to be produced following TBI (Zhang *et al*., 2014).

Although the biological relevance is unclear in TBI, similar autoantibodies have demonstrated pathogenic potential in experimental spinal cord injury (SCI). Injection of sera from mice with SCI into the hippocampi of uninjured mice induced glial activation with prominent neuron loss, whereas sera from B-cell knockout mice had no such effect. Furthermore, these mice developed ectopic meningeal lymphoid follicles, resembling those seen in patients with multiple sclerosis (MS), providing mechanistic insights into how traumatic adaptive immune activation could cause ongoing CNS injury, even after the BBB had been reconstituted after injury (Ankeny *et al*., 2006).

We hypothesised that the release of brain antigens into the systemic circulation and cervical lymph nodes following injury, in the context of a heightened danger-associated molecular pattern (DAMP) milieu, would lead to the generation of autoantibodies against brain proteins. Given the multitude of different brain proteins released, we expected to see autoantibodies generated against a variety of different antigens. To explore the role of autoantibody production in TBI outcome, we developed a CNS human protein microarray, with brain and non-brain antigens, to screen for the development of autoantibodies after moderate to severe TBI, both in the acute phase and at two late time-points. We hypothesised that autoantibodies would develop in the acute phase of TBI, but may persist long-term in some individuals. We lastly hypothesised that these autoantibodies would be pathogenic, and that consequently their presence in the acute phase would associate with worse functional outcome at 6-12 months post-TBI, and their persistence in the longer term would associate with biomarkers of ongoing neurodegeneration.

## Materials and methods

### Study populations

All samples and data were derived from historical observational studies, other than the “Long-term” cohort and healthy controls who were recruited contemporaneously. Sample size was dictated by the availability of samples, and therefore no a priori power calculation was performed. No data was removed from the analysis; all data is displayed in the figures and tables.

Written consent for all patients was obtained from legal representatives prior to enrolment in the acute phase, and further written consent for ongoing use of data and study participation were obtained from the patients at follow up. The studies described were approved either by the Cambridgeshire local research ethics committee (REC 97/290 and 13/EE/0119), or regional ethical board in Stockholm (#2005/1526/31/2).

Healthy controls were recruited through the University of Cambridge (REC 97/290) and Cambridge Biomedical Research Centre (REC 11/33/0007), and all provided written consent. A further bank of 28 “positive control” samples from patients with autoimmune thyroid disease, type 1 diabetes, multiple sclerosis or autoimmune encephalitis provided by Sanja Ugrinovic (Department of Immunology, Addenbrookes Hospital, UK) and Patrick Waters (Nuffield Department of Clinical Neurosciences, University of Oxford, UK) which were used in the early development of the protein microarray contributed normative data.

### Procedures

#### Sample collection and storage

Serum samples were collected at up to four different time-points: 1) “Acute” – within 72 hours of injury (before an adaptive immune response should have occurred), 2) “Subacute – 7-10 days post-injury, 3) “Late” – 6-12 months post-injury, and 4) “Long-term” – 6-13 years post-injury. The samples were aliquoted, labelled with pseudoanonymised identifiers, and frozen immediately at −70°C. Samples from the Validation cohort were shipped on dry ice with temperature monitoring to the University of Cambridge.

#### Demographic, clinical, and outcome information

Demographic information and IMPACT score variables (age, post-resuscitation GCS motor score, pupil reactivity, occurrence of hypoxia or hypotension, Marshall CT classification (Marshall *et al*., 1992), presence of subarachnoid or epidural haemorrhage on CT, blood glucose and haemoglobin concentration (Steyerberg *et al*., 2008)) were recorded by the research team at the time of admission. Overall and extracranial injuries were characterised using the Injury Severity Scale (Baker *et al*., 1974). Glasgow Outcome Score Extended (Jennett and Bond, 1975) was recorded for the Discovery cohort at between 6 and 9 months post-TBI, and Glasgow Outcome Score (Teasdale *et al*., 1998) was recorded at 9-13 months post-TBI, as per the respective original protocols for both of these studies.

#### Autoantibody screening

Autoantibody screening was undertaken using a custom protein microarray based on the HuProt™ (version 2.0) platform (Jeong *et al*., 2012). The custom microarray was devised in collaboration with Cambridge Protein Arrays Ltd. (Cambridge, UK) and CDI laboratories (Puerto Rico) to detect autoantibodies to a broad selection of CNS, BBB and systemic antigens. The custom microarrays consisted of a glass microscope slide with a thin SuperEpoxy coating, printed with quadruplicate spots of recombinant yeast-expressed whole proteins, each fused with GST (glutathione-S-transferase). The array included 79 targets: 52 brain related, 5 blood-brain-barrier related, and 22 non-brain related (full antigen list detailed in **Supplementary Table 1**). Each slide accommodated up to 12 individual serum samples; samples taken at different time-points (Acute, Subacute and Late) from the same patient were apposed on the same slide. The Long-term (6-13 years post-injury) cohort were run in a separate batch alongside healthy controls. In brief, the slides were blocked in 2% BSA/ 0.1% PBS-Tween overnight at 4°C, washed, and then incubated with 200μl of 1:1000 diluted serum at room temperature for 2 hours. The slides were washed again, incubated at room temperature for 2 hours with fluorophore-conjugated goat anti-human IgM-μ chain-Alexa488 (Invitrogen, Carslbad, CA, USA, cat. No. A21215) and goat anti-human IgG-Fc-DyLight550 (Invitrogen cat. No. SA5-10135) secondary antibodies, washed, and then scanned using a Tecan LS400 scanner and GenePix Pro v4 software, with the output being median fluorescence value of the quadruplicate spots for each protein. In order to characterise the reproducibility of the protein microarry, repeat assays were undertaken, both for IgM and IgG responses, in a set of four samples from the Discovery cohort, chosen to reflect the range of autoantibody responses observed. For IgM, the reproducibility of both Subacute polyreactive and antigen specific responses was tested (**see results and Supplementary Fig. 1&2**). For IgG, there were no significant Subacute polyreactive response, and the reproducibility testing focused on the antigen-specific responses.

### Immunoglobulin fraction isolation

To determine whether the microarray results represented true autoantibody responses or non-specific binding related to the acute phase response (Güven *et al*., 2014), the results from serum were compared with purified immunoglobulin fractionated from the same sample. Isolation of both the IgM and IgG fraction of serum was achieved using Kappa and Lambda magnetic beads (PureProteome™). Serum was diluted with PBS (25 µl to a volume of 100 µl) and incubated with 300 µl bead slurry (prepared according to the manufacturer’s instruction) for 60 minutes at room temperature with continuous end-over-end rotation. The beads were separated using a magnetic rack, and the immunodepleted serum removed. The immunoglobulin fraction was eluted from the beads using three washes with 150 µl 1M Glycine HCl, and the resultant eluent was neutralised with 45 µl 1M Tris HCl. Estimation of IgG recovery in the eluent was estimated by NanoDrop™ to be approximately 50%, and thus the eluent was diluted to the equivalent of 1 in 500 before being run as standard on the protein microarray, so as to ensure that the immunoglobulin concentration closely approximated that of 1 in 1000 serum.

### Pre-incubation of serum with cognate antigen

To assess the antigen-specificity of the autoantibodies detected, two serum samples with different autoantibody specificities (anti-MAG IgG and anti-GFAP IgM respectively) were pre-incubated with the corresponding cognate antigen. Serum from each patient was treated in three different ways. Firstly a technical replicate was performed using serum but with no antigen added in order to confirm the original result. Secondly, 200 µl serum was incubated for two hours at room temperature in the presence of excess antigen (5 mcg of MAG or GFAP as appropriate), and lastly 200mcl serum was incubated in the same manner but in the presence of 2.5mcg of both MAG and GFAP to assess the effect of total protein added versus specific cognate antigen. The samples were then diluted to 1 in 1000 (taking into account the volume of solvent from the added protein) and run as standard on the protein microarray.

### Total immunoglobulin quantification

Total IgG and IgM were measured using the standard clinical assay at Addenbrookes Hospital, UK.

### Neurofilament light and glial fibrillary acidic protein quantification

To determine whether the amount of acute antigen released influenced the induction of autoantibody responses, we examined the relationship between acute serum GFAP concentration (defined as the mean serum GFAP concentration over the first 3 days post-injury) and subsequent Subacute anti-GFAP autoantibody level. GFAP is rapidly emerging as the most prominent protein biomarker in TBI (Bazarian *et al*., 2018; Steyerberg *et al*., 2019; Thelin *et al*., 2019; Yue *et al*., 2019), and has been suggested to be the dominant autoantigen following TBI in past reports (Zhang *et al*., 2014).

To assess for ongoing neurodegeneration, serum concentrations of NfL and GFAP were quantified as markers of active brain injury.

Acute GFAP concentrations in the Discovery cohort were measured by Randox Laboratories Ltd (Crumlin, County Antrim, BT29 4QY, United Kingdom), using a sandwich chemiluminescent immunoassay (Evidence Investigator™ Cerebral Custom Array IV). Serum samples were transported on dry ice.

For quantification of NfL and GFAP concentrations in the Validation, Late and Long-term cohorts, serum samples were shipped on dry ice to the Clinical Neurochemistry Laboratory, Sahlgrenska University Hospital where the analyses were performed using commercially available kits (NF-Light Advantage and GFAP Discovery kit, respectively) on the Single molecule array (Simoa) platform, according to instructions from the manufacturer (Quanterix, Billerica, MA, USA)

### Statistical analysis

Descriptive data are presented using median and interquartile range (for continuous variables) or number and percentage (for categorical variables). Between group differences were calculated using Mann-Whitney U tests for unpaired continuous variables and Wilcoxon Matched-Pairs Signed Rank tests for paired continuous variables; complete statistics for all comparisons are tabulated in **Supplementary Table 2**. The association between two continuous variables was assessed using linear regression. Categorical data were compared using chi-squared analysis. Variance of groups was compared using an F-test. Temporal profiles of paired samples were assessed using Friedman with post-hoc Dunn tests. All p-values stated are unadjusted and two-tailed. The Benjamini-Hochberg procedure, with a false discovery rate set to 10%, was used to account for multiple comparisons where appropriate, and p-values which remained <0.05% after correction are highlighted.

### Outcome prediction

The risk of poor outcome was calculated for the acute cohorts using the IMPACT prognostic calculator Core+CT+Lab model (http://www.tbi-impact.org/?p=impact/calc). Where individual covariate data were not available (n = 2% of values; all relating to laboratory results), a value was imputed by using the median value of all patients in that cohort. To assess whether autoantibody responses were related to outcome, we used a sliding dichotomy approach (Murray *et al*., 2005) to group patients according to whether their outcome was “worse than expected” or “better than / as expected” compared with the predictions made by the IMPACT Calculator (**Supplementary Fig. 3**).

### Protein microarray

For each serum sample, the image data were first analysed to extract median fluorescence values of all pixels within each spot. The median fluorescence value of the four quadruplicate spots of each antigen was calculated and used as the value for that antigen in subsequent analysis. Fluorescence values for each antigen were then normalised by dividing by the median value of all antigens for that patient in order to account for any global differences in fluorescence across all antigens that might occur because of technical or biological factors affecting binding affinity.

Both raw (non-normalised) and normalised values were then converted into Z-scores based on respective normal distributions generated from data for each antigen from 269 samples (all samples from the Discovery and Validation cohorts, healthy controls and “positive” controls); the top and bottom 2% of values for each antigen were excluded to remove artefactual outliers or strong positive results so that the interpretation of Z-scores approximates those of a Gaussian distribution. This broad selection of subjects included both “well” and critically ill participants to ensure it would include variation attributable to non-specific binding resulting from acute phase reactants (Güven *et al*., 2014). While this approach may have reduced sensitivity to detection of antibody responses, we accepted this as a price for the increased specificity that resulted. A representative distribution of Z-scores within all samples assayed is displayed in **Supplementary Fig. 4**. For the initial screen, a positive autoantibody result was defined by a threshold of Z≥3. An increase of Z≥1 between paired samples, providing the second sample showed a Z ≥3, was used to define the development of a new autoantibody.

Analyses were performed using Graphpad Prism (version 8.1.0; Graphpad Software) and SPSS (version 25; IBM SPSS).

### Data availability

All data used in this study are available upon request from the corresponding author.

## Results

### Study Populations and measurements

Banked samples of serum collected at up to two time-points during the first week of admission (“Acute”: 0-3 days post-TBI, and “Subacute”: 7-10 days post-TBI) from two retrospective cohorts of patients with moderate to severe TBI, recruited from two separate centres, were studied sequentially as Discovery and Validation cohorts. The Discovery cohort (n=25) was recruited from patients admitted to the Neurosciences Critical Care Unit, Addenbrookes Hospital, UK between January 2012 and August 2013, and the Validation cohort (n=66) was recruited from patients admitted to the Neurointensive Care Unit, Karolinska University Hospital, Sweden between January 2007 and October 2012. The demographic details of both acute cohorts are summarised in **Table 1**. The Validation cohort were older (p=0.001), more likely to have a mass lesion (p=0.001) and traumatic subarachnoid haemorrhage (p=0.001), but had lower overall trauma severity (based on the injury severity scale (ISS); p=0.0003). They had a poorer predicted outcome based on the IMPACT variables (p<0.0001), but Glasgow Outcome Score (GOS) at follow up was not different (p=0.125). The Validation cohort was followed up at a median of 12 months post-injury compared with 7 months for the Discovery group.

**Table 1.**
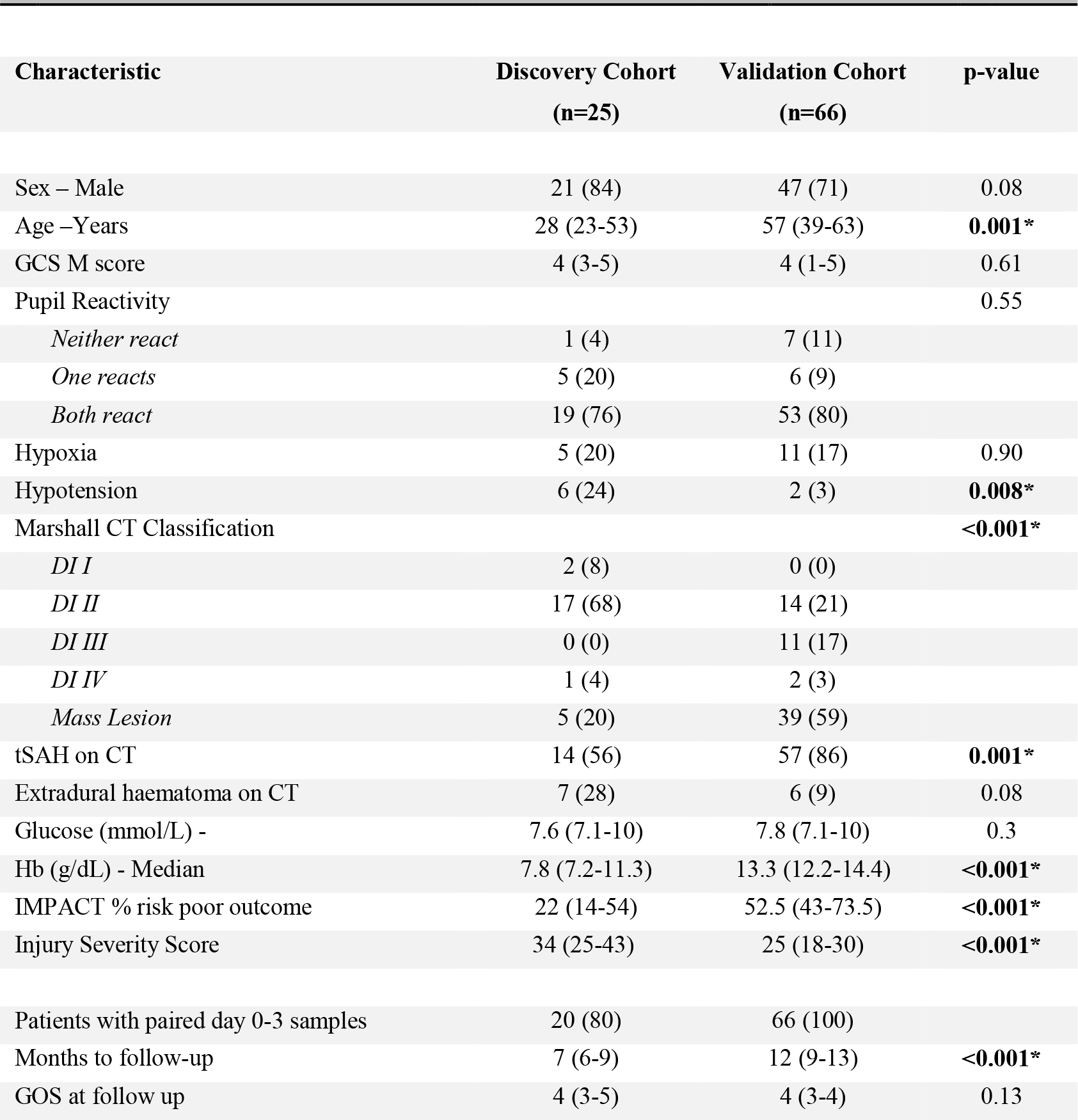
Baseline Characteristics of Acute TBI Cohorts. Baseline characteristics of the Discovery and Validation cohorts. *GCS = Glasgow Coma Scale, DI = Diffuse injury, tSAH = traumatic subarachnoid haemorrhage*., *GOS = Glasgow Outcome Score. Categorical data are presented as Number (%), continuous variables as Median (IQR). * denotes p-values which remain significant following the Benjamini-Hochberg procedure*.

| Characteristic | Discovery Cohort<br>(n=25) | Validation Cohort<br>(n=66) | p-value |
| --- | --- | --- | --- |
| Sex – Male | 21 (84) | 47 (71) | 0.08 |
| Age –Years | 28 (23-53) | 57 (39-63) | **0.001\*** |
| GCS M score | 4 (3-5) | 4 (1-5) | 0.61 |
| Pupil Reactivity |  |  | 0.55 |
| *Neither react* | 1 (4) | 7 (11) |  |
| *One reacts* | 5 (20) | 6 (9) |  |
| *Both react* | 19 (76) | 53 (80) |  |
| Hypoxia | 5 (20) | 11 (17) | 0.90 |
| Hypotension | 6 (24) | 2 (3) | **0.008\*** |
| Marshall CT Classification |  |  | **<0.001\*** |
| *DI I* | 2 (8) | 0 (0) |  |
| *DI II* | 17 (68) | 14 (21) |  |
| *DI III* | 0 (0) | 11 (17) |  |
| *DI IV* | 1 (4) | 2 (3) |  |
| *Mass Lesion* | 5 (20) | 39 (59) |  |
| tSAH on CT | 14 (56) | 57 (86) | **0.001\*** |
| Extradural haematoma on CT | 7 (28) | 6 (9) | 0.08 |
| Glucose (mmol/L) - | 7.6 (7.1-10) | 7.8 (7.1-10) | 0.3 |
| Hb (g/dL) - Median | 7.8 (7.2-11.3) | 13.3 (12.2-14.4) | **<0.001\*** |
| IMPACT % risk poor outcome | 22 (14-54) | 52.5 (43-73.5) | **<0.001\*** |
| Injury Severity Score | 34 (25-43) | 25 (18-30) | **<0.001\*** |
| Patients with paired day 0-3 samples | 20 (80) | 66 (100) |  |
| Months to follow-up | 7 (6-9) | 12 (9-13) | **<0.001\*** |
| GOS at follow up | 4 (3-5) | 4 (3-4) | 0.13 |

A subset of the Discovery cohort (n=20) provided serum samples 6-12 months post-injury, forming a third “Late” cohort. A fourth “Long-term” cohort (n=34, 13 of whom had contributed to the Discovery cohort, and six of whom had samples at all four time-points) provided new serum samples 6 to 13 years post-TBI.

Healthy controls (n=45) were recruited through the University of Cambridge and Cambridge Biomedical Research Centre (demographic comparisons with relevant cohorts shown in **Supplementary Table 3**).

### A polyantigenic IgM response of variable magnitude is seen 7 days post-TBI in most individuals from the Discovery cohort

In the 20 patients in the Discovery cohort with paired Acute (day 0-3 post-TBI) and Subacute (day 7 post-TBI) samples, an upregulation in IgM antibodies to most antigens was seen in a group-wise comparison between the Acute and Subacute (p<0.0001) time-points, as was a similar but smaller increase in IgG (p=0.035) (**Fig. 1A; Supplementary Table 2**). The spread of the median Z-score of all IgM responses to antigens for each patient was greater for the Subacute than the Acute cohort (F=0.409, p=0.004; **Fig. 1C**), suggesting that the later response varied more between individuals. Minimal inter-individual variation was seen in IgG response (F=1.16, p=0.75; **Fig. 1D**). Repeat assays in a set of four samples, spanning the range of broad polyantigenic IgM responses seen, showed good reproducibility, with identical rank order for fluorescence. The Subacute IgG responses showed little variation between patients, and therefore comparison of rank orders was not appropriate. Non-specific binding mediated by other factors such as acute phase reactants (e.g. CRP) was excluded by demonstrating that purified immunoglobulin (Ig) from sera showed the same results as unfractionated sera (**Supplementary Fig. 1A**). There was no relationship between the IgG binding in serum and purified Ig fraction, in keeping with the negligible inter-individual variation seen (**Supplementary Fig. 1B**).

**Figure 1.**
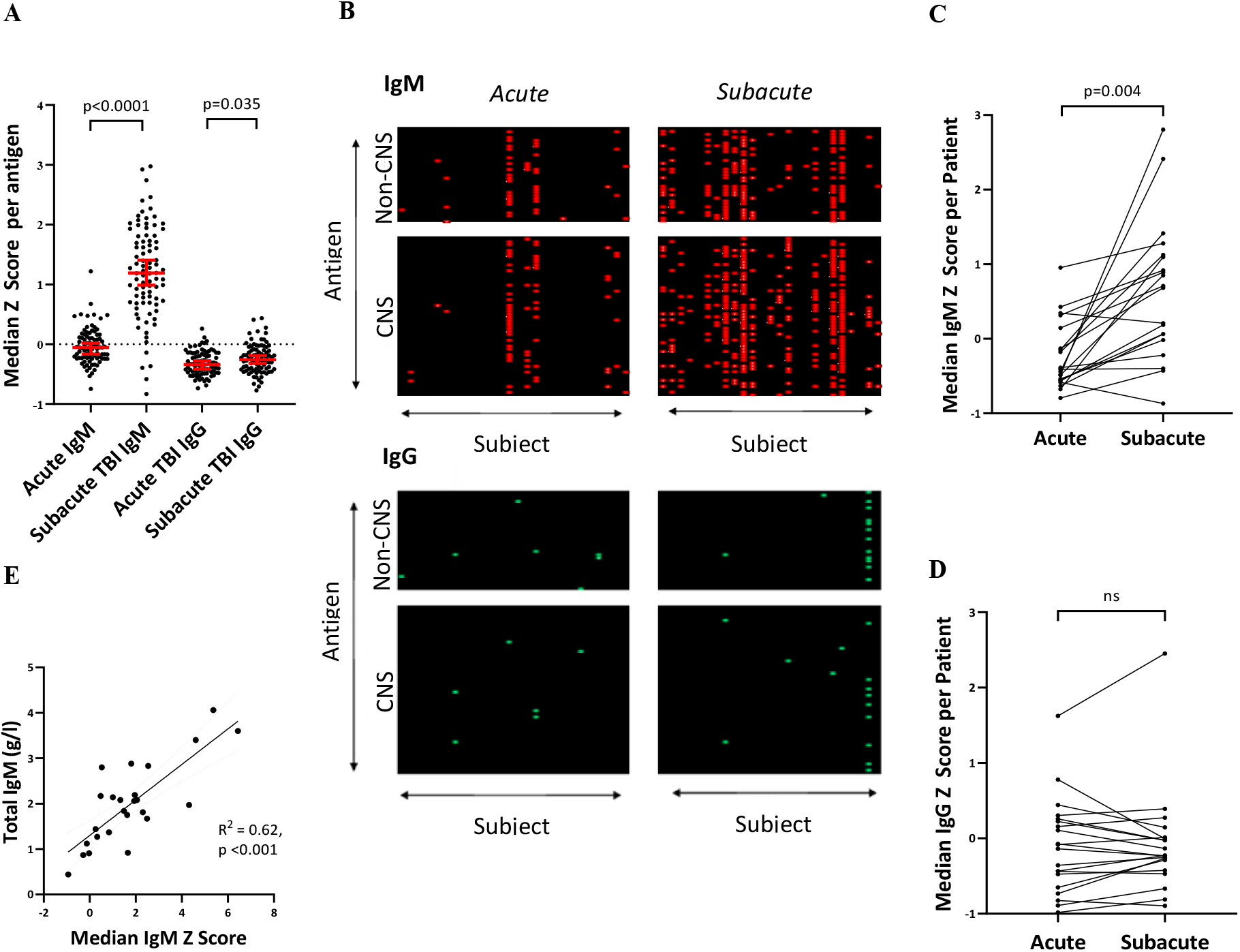
(**A**) Marked increase in polyantigenic IgM and smaller increase in polyantigenic IgG is seen between the Acute and Subacute time points at a group-level in the 20 patients with paired serum from the Discovery cohort. Each datapoint refers to the median Z-score of an antigen across the cohort. (**B**) Heatmaps, where antigens Z≥3 are highlighted, display the polyantigenic nature of IgM, and to a lesser degree, IgG, responses. (**C&D**) The median Z score of all antigens per patient captures the degree of inter-individual variation of IgM, and comparatively homogenous IgG, polyantigenic responses. (**E**) Total serum IgM concentration correlated with an individual’s median IgM Z-score derived from the protein microarray. *Statistical tests for A: Wilcoxon Matched-Pairs Signed Rank test; C&D: F-test from one-way ANOVA E: Linear regression*.

Total serum IgM concentration, measured using a standard clinical assay, was increased in half of patients (**Supplementary Fig. 1C**), and correlated with the median IgM Z-score derived from the protein microarray (R^2^=0.62, p<0.001) (**Fig. 1E**). Serum IgG concentrations, conversely, were reduced, with 13/25 (52%) of patients’ values below normal reference levels (**Supplementary Fig. 1D**). Unlike IgM, there was minimal spread between patients, suggesting that reduction in IgG was a more homogenous phenomenon. There was no relationship between total serum IgG and mean IgG Z-score on the protein microarray (**Supplementary Fig. 1E**).

### Dominant responses to specific antigens are seen in the majority of individuals from the Discovery Cohort

To identify whether there were any dominant autoantibody responses to specific antigens that stood apart from the global upregulation, the data were normalised (see methods) and antigen responses with high Z-scores identified.

Five of 20 patients in the Discovery cohort (25%) developed new IgM autoantibodies and 13/20 (65%) patients developed new IgG autoantibodies between the Acute and Subacute sampling points (**Fig. 2A**). These findings were verified using the purified Ig fraction (**Supplementary Fig. 2A**) and the antigen-specificity of these responses demonstrated by the attenuation of the response when serum was pre-incubated with the antigen of interest (**Supplementary Fig. 2B**).

**Figure 2.**
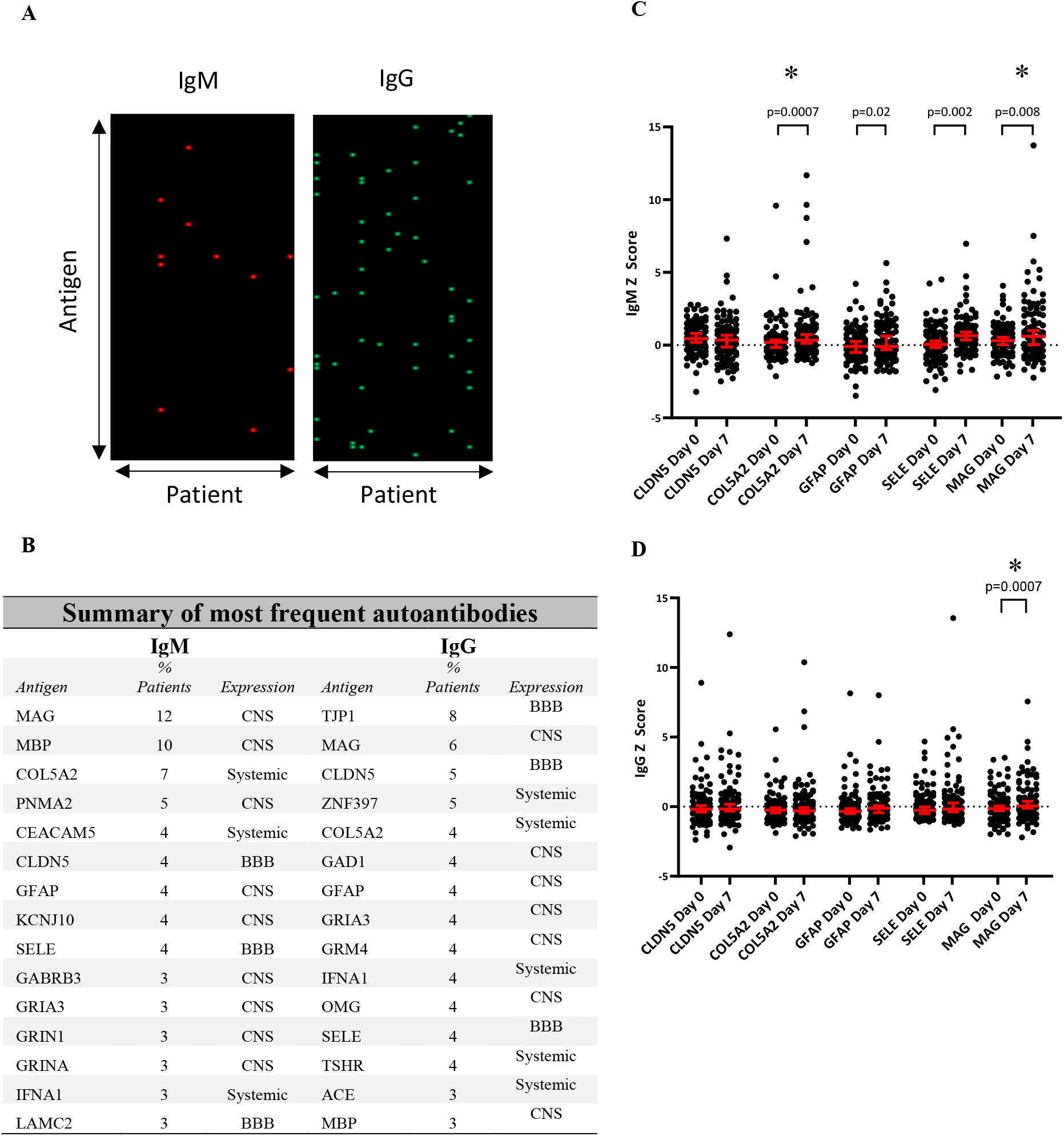
(**A**) Heatmaps displaying dominant IgM and IgG autoantibodies to specific antigens newly detected at 7-10 days post-injury (defined as Z>3 with a Z increase of >1 from the paired Acute sample). Each row corresponds to an antigen, and each column to an individual patient. (**B**) Top 15 most frequent autoantibodies for both IgM and IgG isotypes seen following TBI at the Subacute time point across both Discovery and Validation cohorts. “Expression” refers to the predominant site of expression. CNS = Central nervous system; BBB = Blood brain barrier (**C&D**) Comparison of whole group IgM and IgG Z score change between Acute and Subacute samples for the five most frequently seen autoantibodies. *Statistical test: Wilcoxon Matched-Pairs Signed Rank test; * denotes comparisons which remain significant even after removal of all values where Z≥3*.

At the Subacute time point, antigens with high IgG responses were almost ten times more likely to also have an IgM response compared to antigens with a low IgG response (5/16 [31%] vs. 25/568 [4.4%], OR 9.87, p=0.0001). In contrast, at the early time point, there was no such relationship (1/11) [9%] vs 20/538 [2.6%], OR=2.52, p = 0.37).

### The Discovery cohort results are replicated in the larger Validation cohort from a different centre

The findings from the Discovery cohort were tested in the Validation cohort. The increase in polyantigenic IgM and IgG seen between the Acute and Subacute samples were replicated (p<0.001 and p=0.003, respectively; **Supplementary Table 2**). The development of dominant IgM and IgG responses to specific antigens, after normalisation, were seen in 42/66 (64%) and 37/66 (56%) patients respectively.

### Subacute autoantibody responses are associated with functional outcome at 6-12 months post-injury in the Discovery cohort

The data from the Discovery and Validation cohorts were pooled to investigate whether the autoantibody responses correlated with clinical or demographic parameters. Young age was associated with a high polyantigenic IgM response (R^2^=0.19, p<0.0001), but none of the following parameters correlated with either IgM or IgG responses: Sex, Glasgow Coma Scale Motor Score, Injury Severity Score and IMPACT score. The older age of the Validation cohort meant that there were fewer patients with a high magnitude of polyantigenic IgM response (**Supplementary Table 2**).

Given the notable variation in autoantibody responses between individuals, we sought to investigate whether these responses associated with outcome. Patients were dichotomised by outcome depending on whether their actual functional outcome at 6-12 months was as good as/better than predicted by the IMPACT prognostic model, or worse (**see Materials and Methods and Supplementary Fig. 3A**), and the autoantibody profiles were compared between groups. In the 25 patients from the Discovery cohort, those with an outcome worse than predicted had a higher median IgM Z-score than those whose outcome was as good as, or better than expected (p=0.01) (**Fig. 3A; Supplementary Table 2**), and indeed the median IgM Z-score provided moderate discriminatory power to differentiate the two groups (AUC 0.776, p=0.019; **Fig. 3B**). There was no difference between groups in the number of dominant IgM or IgG antibodies to specific antigens (p=0.70).

**Figure 3.**
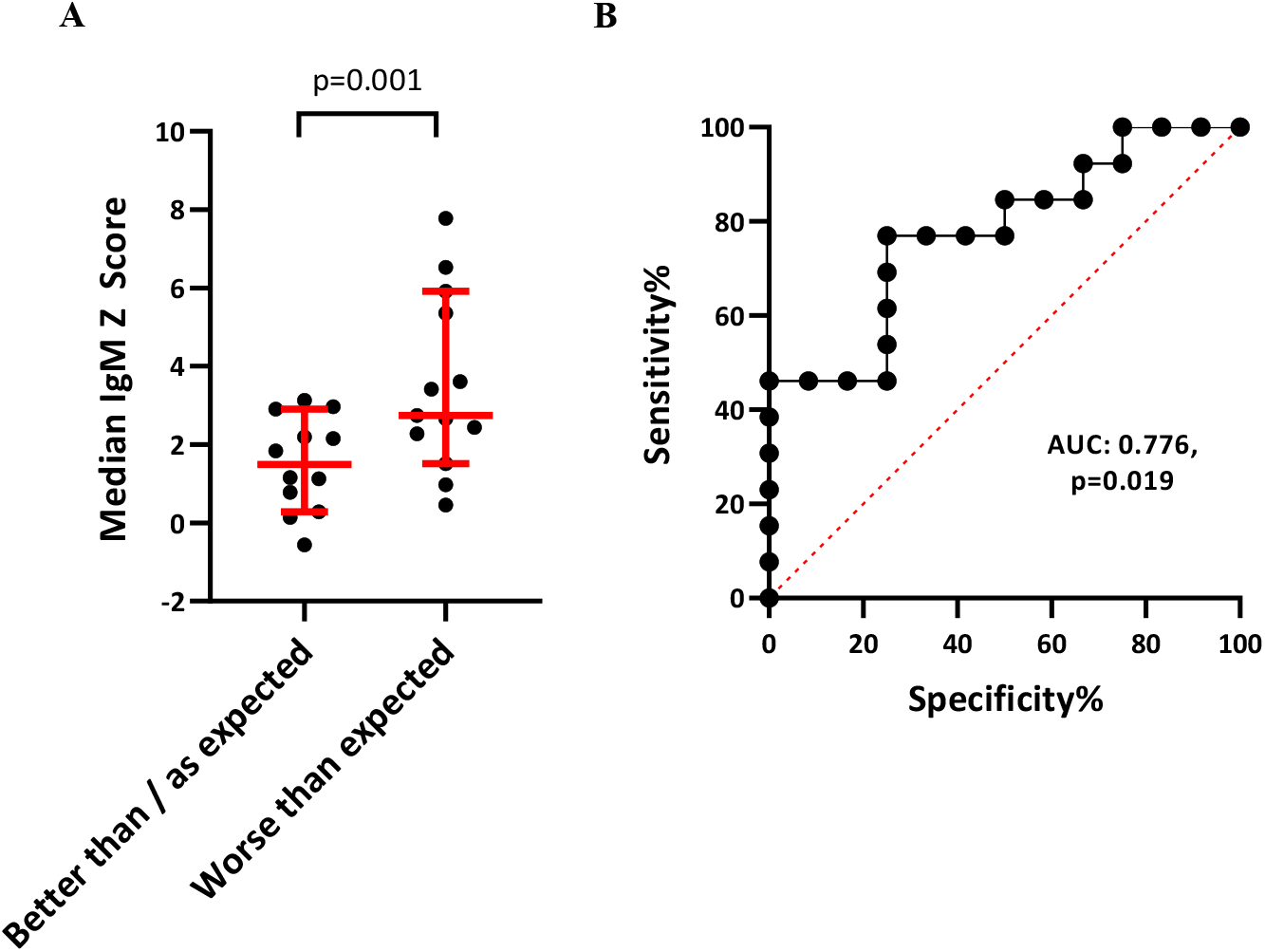
(**A**) Comparison of polyantigenic IgM response between the dichotomised prognostic groups in the Discovery cohort suggests that more marked responses are associated with an outcome worse than predicted by the IMPACT variables. *Statistical test: Mann-Whitney U test*. (**B)** ROC curve analysis displaying the ability of polyantigenic IgM response to differentiate between “Better than / as expected” vs. “Worse than expected” prognosis groups.

The same approach was used in the Validation cohort, although the granularity was less as only GOS had been collected (**Supplementary Fig. 3B**), and the outcome data had been gathered at a median of twelve months rather than seven. In this cohort, no relationship between any autoantibody profile and outcome was demonstrable (median IgM Z p=0.15; number of normalised IgM antibodies Z≥3 p=0.67; number of normalised IgG antibodies Z≥3 p=0.33) (**Supplementary Table 2**).

### Dominant responses occurred most frequently to certain antigens

The median normalised data for all patients with paired Acute and Subacute samples from both Discovery and Validation cohorts were assessed for the most frequent autoantibodies. Autoantibodies were seen to 51 different antigens in total, with 44 different targets for IgM autoantibodies, and 31 for IgG. Of these, 24/51 (47%) antigens induced both IgM and IgG responses. The 15 most common targets for each isotype are displayed in **Fig 2B**. The most frequently seen overall targets were MAG, COL5a2, CLDN5, GFAP and SELE. Of note, the autoantibodies did not solely target CNS antigens. While CNS targets comprised most of the 15 most commonly seen autoantibodies (9/15 IgM, 7/15 IgG), autoantibodies to BBB antigens (TJP1, CLDN5 and SELE), were also common (3/15 for both IgM and IgG, potentially the most common autoantibody group proportionately given the comparatively few BBB targets on the protein microarray [5 BBB targets compared with 52 CNS and 20 systemic]). There were also autoantibodies to systemic antigens (3/15 IgM, 5/15 IgG); for instance we commonly observed responses against COL5a2, a ubiquitous collagen, autoantibodies against which are recognised in respiratory disease and implicated in rejection of lung transplants (Iwata *et al*., 2008; Tiriveedhi *et al*., 2013). In the Discovery group (where trauma computerised tomography series reports were available), the Subacute IgG response to COL5a2 was higher in patients with lung contusions than those without (p=0.04, **Supplementary Table 2**).

Our use of a reference distribution that included patient data, and the use of a threshold of Z≥3 represented a very stringent basis for detecting autoantibody responses. Consequently, to assess whether the use of this process was hiding a broader group effect, the Z-scores for the five most frequently seen autoantibodies were compared between Acute and Subacute samples for every patient. 4/5 IgM responses (against COL5a2, GFAP, SELE and MAG) and 1/5 IgG responses (against MAG) showed a group difference (**Fig. 2C&D, Supplementary Table 4A-D**). Even when high results (Z≥3) were removed, the differences in IgM responses to COL5a2 and MAG, and the IgG response to MAG, remained significant. These analyses suggest that thresholding at a specific Z-score may have underestimated the proportion producing autoantibodies to (at least) these proteins.

To assess the relationship between acute protein release and subsequent autoantibody response, the acute serum GFAP concentration was compared with Subacute GFAP autoantibody responses. Given that serum GFAP concentration for the Discovery and Validation cohorts had been measured using different platforms, the cohorts were analysed separately. A significant relationship was seen between GFAP levels and anti-GFAP IgG Z-scores in the Discovery cohort (Spearman rho 0.58; p=0.008). These findings were not replicated in the Validation cohort however (Spearman rho 0.19; p=0.12).

### Polyantigenic autoantibody responses persist for several years post-injury

A subset of 20 patients from the Discovery cohort provided samples at a Late time-point (6-12 months post-TBI, **Supplementary Table 3**). Late polyantigenic responses (as measured by non-normalized Z-scores) were significantly higher for both IgM and IgG than controls (p <0.0001 for both; **Supplementary Table 2**), and than Acute levels in a within-subject comparison (p<0.0002 for IgM and p<0.0001 for IgG, respectively; **Fig. 4A, Supplementary Table 2**).

**Figure 4.**
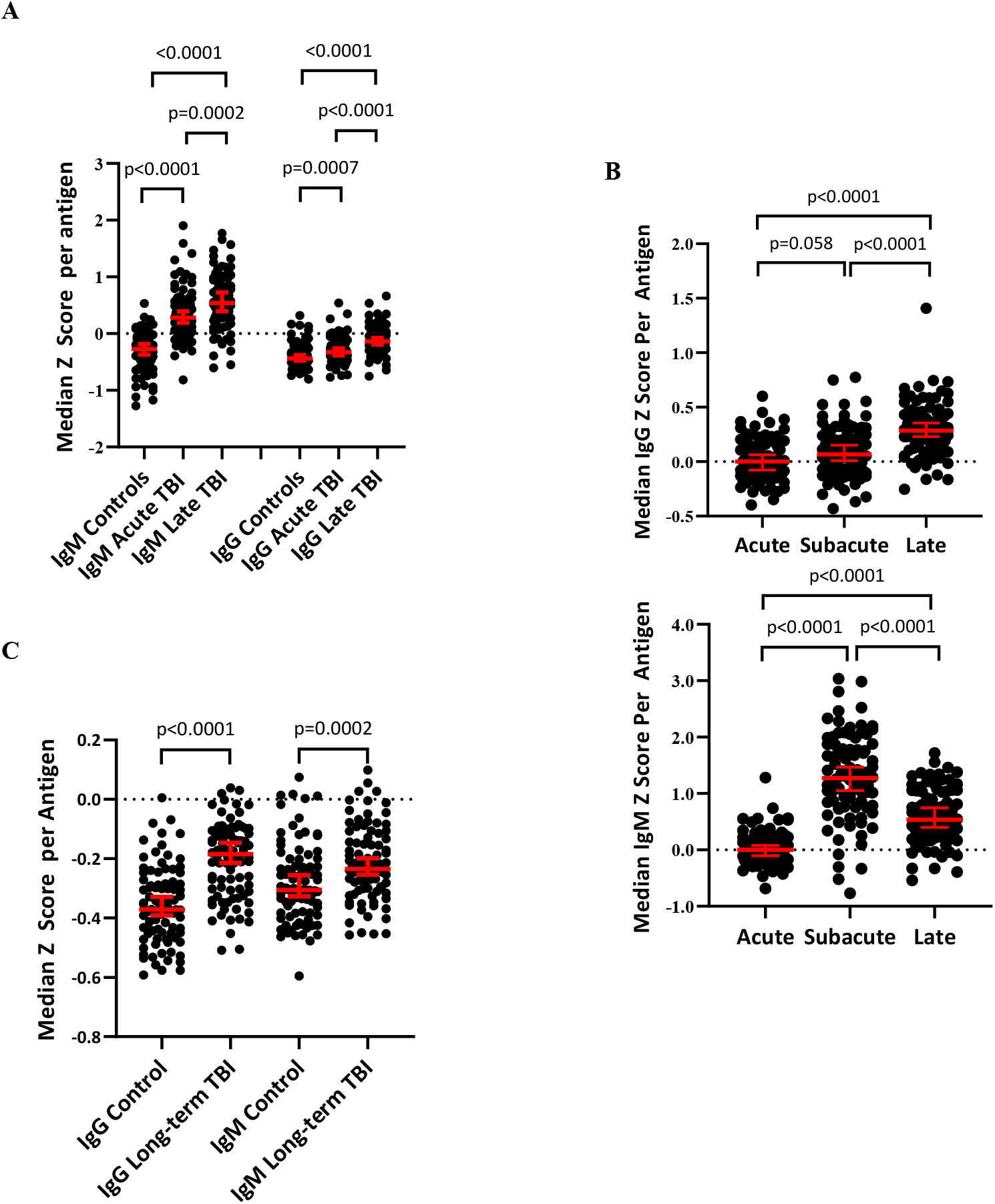
(**A**) A Late increase in polyantigenic IgM and IgG is seen in patients 6-12 months post-TBI compared with their paired Acute samples. Both samples show higher reactivity than healthy controls. (**B**) Temporal profiling of polyantigenic autoantibody responses display Subacute IgM and Late IgG peaks. (**C**) Long-term persistence of (particularly IgG) polyantigenic responses are seen in patients 6-13 years post-injury compared with healthy controls. *Statistical tests for paired TBI samples: Wilcoxon Matched-Pairs Signed Rank test; for control vs. TBI: Mann-Whitney U test*.

A more complete temporal analysis of data from Acute, Subacute, and Late samples revealed an intriguing difference between the pattern of IgM and IgG responses. IgM reactivity peaked at the Subacute time-point before falling towards (but not back to) baseline, whereas IgG reactivity was maximal at the Late time-point (p<0.0001 for all comparisons except Acute vs. Subacute IgG where p=0.058; **Fig. 4B)**.

A further Long-Term cohort who had sustained a moderate to severe TBI between 6 and 13 years previously (n=34, of whom 13 had contributed to the Discovery cohort) were screened for autoantibodies, and showed persistent polyantigenic IgM and (particularly) IgG responses compared to healthy controls (p=0.0002 and p<0.0001 for IgM and IgG, respectively; **Fig. 4C, Supplementary Table 2**). As this cohort was processed separately to the Discovery cohort, the non-normalised data could not be directly compared with matching samples due to the risk of batch effects. The 28 age and sex-matched healthy controls used in this analysis were therefore recruited separately.

### Dominant autoantibody responses to specific antigens persist for at least a year post-TBI but have waned by 6-13 years post-injury

To determine whether there was a persistence of dominant autoantibody responses to specific antigens, the five most frequently detected autoantibodies seen during the acute-phase were assessed in the Late samples after normalisation. There was indeed a persistence of IgG against MAG and SELE, but also interestingly an IgM to MAG (**Fig. 5A&B; Supplementary Tables 5&6**).

**Figure 5.**
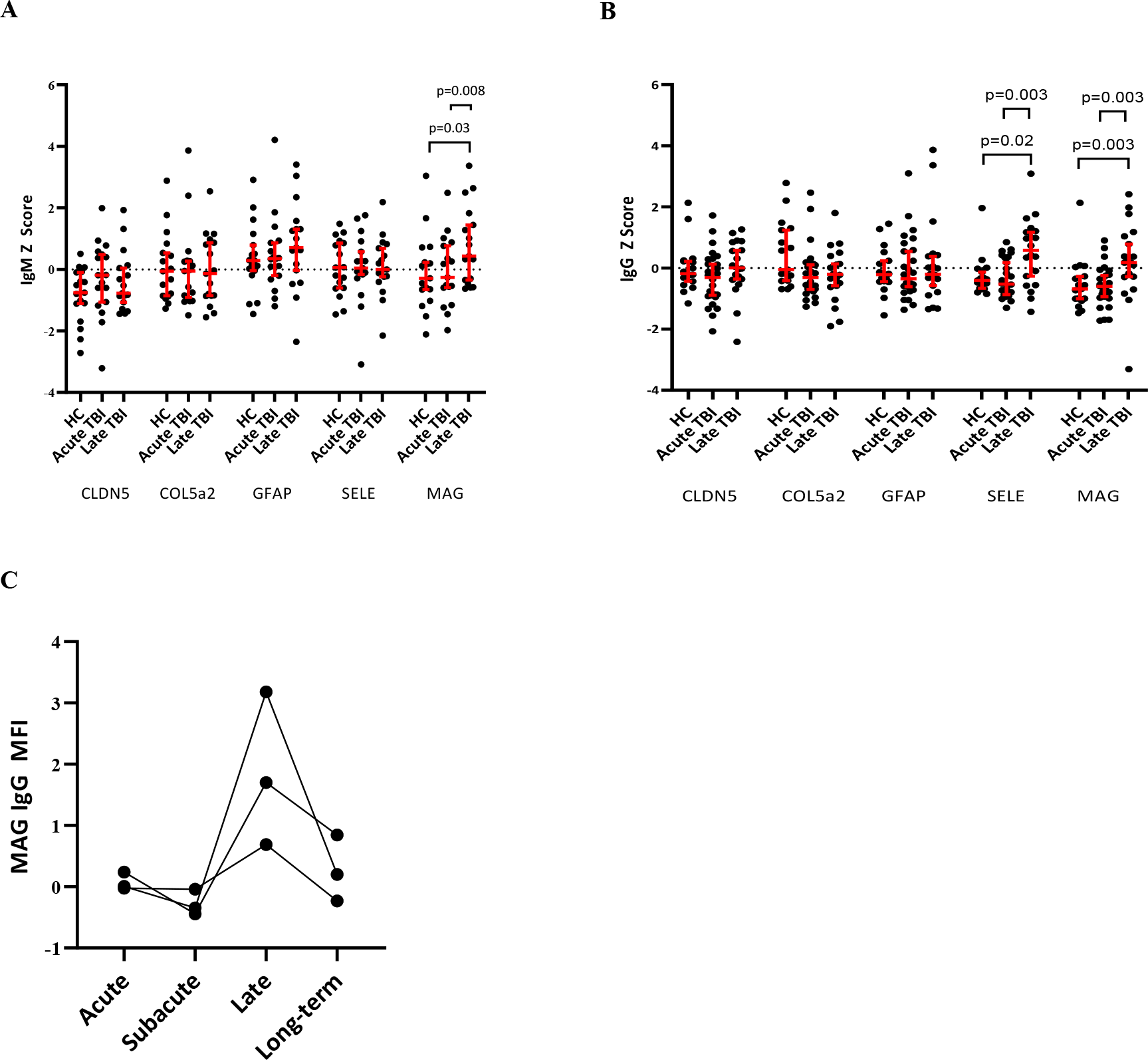
(A**&B**) Screening for the five most frequently seen autoantibodies in the acute phase reveals Late persistent IgM to MAG and IgG to MAG and SELE. *HC = healthy controls* (**C**) The temporal profiles of IgG autoantibodies from three patients with serum taken at 4 time-points who developed anti-MAG IgG antibodies at Late time-points (6-9 months) showing a return towards baseline Long-term (7-13 years). *Statistical tests for paired TBI samples: Wilcoxon Matched-Pairs Signed Rank test; for control vs. TBI: Mann-Whitney U test*.

To assess whether Late IgG autoantibodies were the result of Subacute class-switching, the data from the 11 patients with samples taken at Acute, Subacute and Late time-points were analysed for temporal relationships. Those antigens where a Late IgG Z≥3 was seen were more likely to have a corresponding Subacute IgM of Z≥2 (this lower threshold again used to capture more subtle responses) than those antigens where Late IgG Z<3 (7/27 [26%] vs. 58/919 [6%], OR 5.2, p<0.0001). There was no such relationship seen between Late IgG and Acute IgM (1/27 [4%] vs. 38/919 [4%], OR= 0.89, p=0.91, suggesting that the Late IgG are indeed related to Subacute IgM production.

Persistence of these antibodies was not seen in the Long-term samples however (p>0.05 for all comparisons with controls at this time-point; **Supplementary Table 2**), suggesting that these dominant responses had waned over the years between the Late and Long-term samples (**Fig. 5C**).

### Markers of neurodegeneration are seen in a subset of patients years after TBI, and differ depending on autoantibody profile

In order to investigate the relationship between the persistence of autoantibodies and progressive neurodegeneration following TBI, serum concentrations of NfL and GFAP were measured and compared with the median non-normalised IgG and IgM Z-score of each patient (representing the polyantigenic response) and the top four most frequently seen dominant responses to specific CNS antigens detected following normalisation.

Both NfL and GFAP concentrations were significantly higher at a group-level in the Late TBI (6-12 months post-injury) cohort than healthy controls (NfL p<0.0001; GFAP p=0.05; **Fig. 6A; Supplementary Table 2**), and there was an association between the concentrations of the two proteins (R^2^=0.3, p=0.002); **Fig. 6B)**. Although the effective half-life of NfL in the serum has not been fully delineated (but believed to be in the region of a few weeks), the effective half-life of GFAP is between 24-48 hours (Thelin *et al*., 2017), and thus the presence of raised GFAP levels, whilst less marked, is highly suggestive of an active injury process, rather than slow clearing of protein released at the time of injury. Whilst at the Long-term (6-13 year) time-point there was no significant group difference in serum GFAP or NfL concentration between TBI patients and healthy controls (GFAP p=0.11, NfL p=0.4; **Fig. 6C, Supplementary Table 2**), a larger proportion of TBI patients had neural injury biomarkers above the control normal range (defined as values within 2 standard deviations of the control population mean) (GFAP 7 [20%] vs. 1 [0.25%], p=0.01; NfL 7 [20%] vs. 2 [0.5%], p=0.046), suggesting that at least a subset of patients experience ongoing neurodegeneration, in keeping with previous studies (Ruff *et al*., 1991; Millis *et al*., 2001; Sander *et al*., 2001; Whitnall, 2006; Till *et al*., 2008; Newcombe *et al*., 2016).

**Figure 6.**
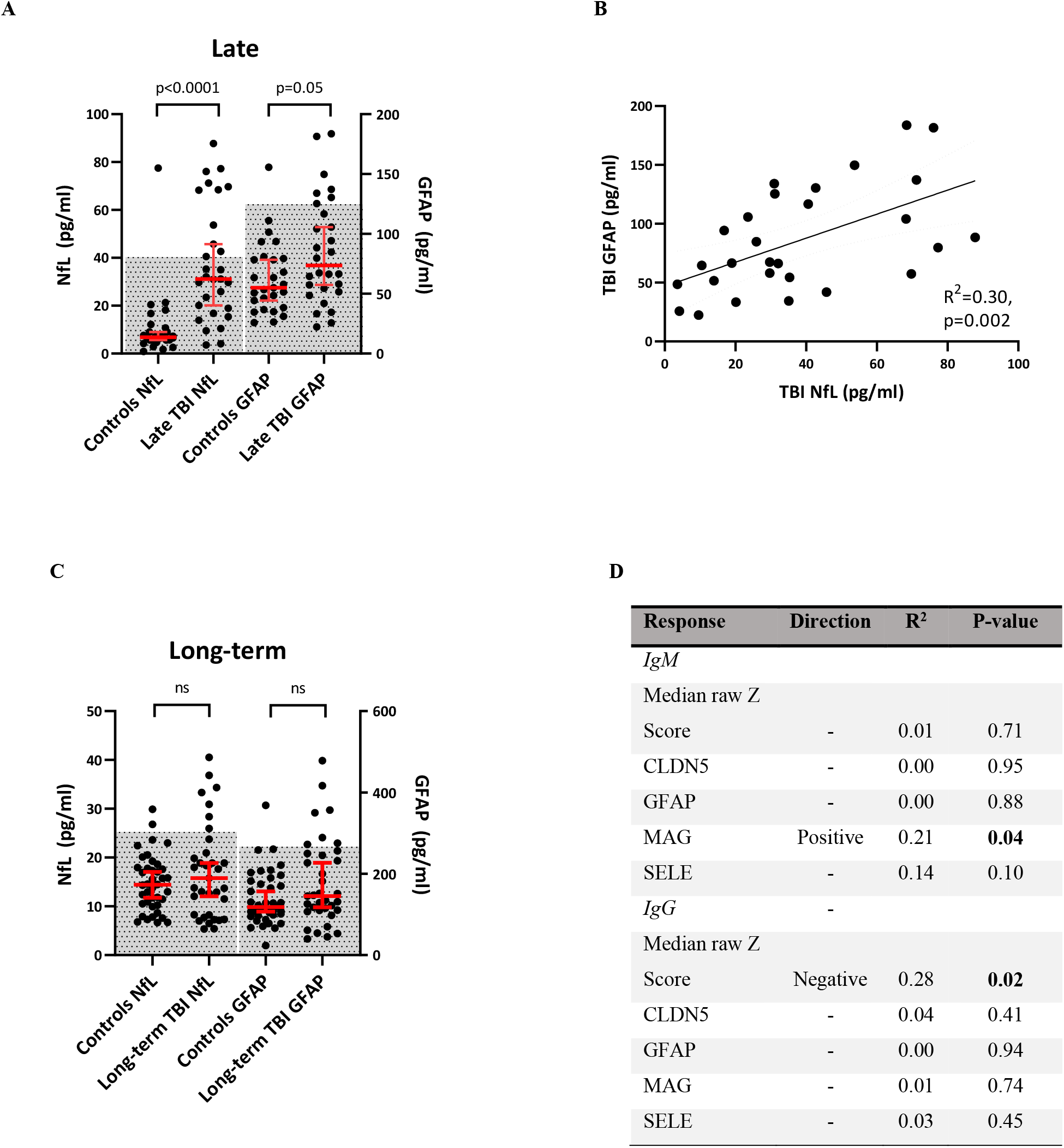
(**A**) Late TBI patients have higher serum NfL and GFAP concentrations than healthy controls at a group level (*hatched area = mean +/- 2 standard deviations*) (**B**) Late serum NfL and GFAP concentrations covary (**C**) There is no difference at a group level between Long-term TBI patients and healthy controls, but more TBI patients lie outside the normal range (*hatched area = mean +/- 2 standard deviations)*. (**D**) Anti-MAG IgM weakly correlates with high serum NfL concentrations, whereas polyantigenic IgG antibodies correlate with low serum NfL concentrations. *Statistical tests for A&C:Mann-Whitney U test; for B&D: Linear regression*.

As NfL appeared to be the more sensitive of the two biomarkers to discriminate from healthy controls, the Late autoantibody profiles were regressed against serum NfL concentrations. Although no association survived adjustment for multiple comparisons, two hypotheses were generated by this analysis: 1) high anti-MAG IgM reactivity associates with high NfL, and 2) polyantigenic IgG associates with low NfL (**Fig. 6D**). The polyantigenic IgG association was not present in the Long-term cohort; there was again a suggestion that high anti-MAG IgM weakly correlated with high NfL (R^2^=0.15, p=0.029), but this relationship was largely driven by a single individual.

## Discussion

In this study, we identified the presence of two discrete autoantibody responses following TBI. The first is a “polyantigenic” increase in IgM (and less prominently IgG) responses against many antigens, commencing within the first week of TBI; the second comprises clear dominant responses to a small number of neural antigens, of which MAG was the most common, but also to blood-brain-barrier antigens (TJP1, CLDN5) and systemic antigens (such as COL5a2). The temporal profile of these responses largely recapitulated a typical primary adaptive immune response, with early IgM and later IgG production. The Late (6-12 months) and Long-term (up to 13 years) persistence of widespread IgM autoantibody responses, however, is more in keeping with an alteration in “natural antibody” repertoire, which are (by their nature) low-affinity polyreactive species (Palma *et al*., 2018).

The polyantigenic IgM response was less prominent with increasing age, in keeping with the known effect of ageing on both B-1 (polyreactive IgM) and B-2 (antigen-specific) responses (Sasaki *et al*., 2011; Rodriguez-Zhurbenko *et al*., 2019). In younger patients (where they were most prominent), this response appeared to account for a proportion of the variance between actual clinical outcome and the outcome predicted by the IMPACT variables, and did not seem to be a surrogate of other clinical markers such as injury severity score.

An association between autoantibody production and late functional outcome was also suggested by the observation that protein biomarkers of ongoing neural injury were persistently elevated at a group level at 6-12 months after injury, at which time-point their levels correlated with levels of anti-MAG IgM autoantibodies. While the protein biomarker levels were less consistent 6-13 years post-TBI (with no significant elevation compared to controls at the group level), they remained elevated in a subset of patients, and again correlated positively with the presence of anti-MAG IgM autoantibodies. The primacy of MAG as an autoantigen at late time-points is particularly interesting given that white matter tract degeneration is a key phenomenon driving late post-traumatic neurodegeneration (Sullivan *et al*., 2013; Scott *et al*., 2015; Newcombe *et al*., 2016). Intriguingly, NfL levels at 6-12 months correlated inversely with levels of polyantigenic IgG autoantibodies, which would be in keeping with the putative role of these antibodies as homeostatic agents playing an active role against ongoing detrimental processes driving neurodegeneration (Palma *et al*., 2018); indeed natural IgG antibodies have been shown to block the neurotoxic effect of amyloid-beta in mouse models of Alzheimer’s disease (Dodel *et al*., 2011).

We explored the contribution of antigen exposure as a potential driver of autoantibody production. A recent study revealed that GFAP and NfL serum levels measured the first week following TBI, together with a panel of other commonly used protein biomarkers of brain injury, revealed the strongest association to long-term outcome (Thelin *et al*., 2019). In our study, acute levels of GFAP (measured as a biomarker of brain injury) were related to subsequent levels of cognate autoantibodies in one cohort of patients, but this was not replicable. It is possible that the level of antigen exposure in the acute phase is a necessary (but not sufficient) modulator of autoantibody production, but further work is needed to address this issue. In any case, clear separation of the influence on the autoantibody response would require a larger sample of patients, since higher protein levels may not just represent a higher acute antigen exposure, but also indicate a more severe injury, which may in turn generate a more robust host response.

These results raise important questions regarding the mechanisms, range, and impact of autoantibody production following TBI. While it might be hypothesised that the dominant responses to specific antigens result from self-antigen release in the context of upregulated DAMP signals in a vaccination-type manner, the biology of the ongoing polyantigenic responses is less clear. The concept of “naturally-occurring” autoantibodies, which are thought to play a homeostatic role including the clearance of apoptotic cell debris (Grönwall *et al*., 2012; Palma *et al*., 2018), is well recognised although poorly understood, and resonates with our findings. The classical description of these autoantibodies is of low-affinity polyreactive IgM species which are produced by B1 cells following T-independent activation (Grönwall *et al*., 2012). The existence of natural IgG autoantibodies has also been mooted, but the literature is even more sparse (Panda *et al*., 2013). That such a response might occur acutely in response to massive cell injury is easy to appreciate, but the ongoing alteration of this system years after the injury is surprising, and its implications as yet unknown.

The finding of dominant autoantibodies against multiple antigens developing in single individuals after TBI is in keeping with previous western blot experiments (Zhang *et al*., 2014), but the most frequent target seen differs (MAG vs. GFAP). This difference may perhaps simply be explained by the relative capabilities of the two assays to detect a particular autoantibody, but the effect of the polyantigenic autoantibody responses may be a complicating factor that might be detected by the platforms in different manners. Indeed, the prominence of the polyantigenic autoantibody response has significant implications on the further study of this field. Any autoantibody assay (such as ELISA or cell-based assay) is likely to be significantly affected by this response, and thus might falsely report an antigen-specific response. However, although a protein microarray approach allows for detection of the polyantigenic response, the sensitivity of protein microarrays for any given autoantibody may inherently be lower than an assay honed for the detection of a specific autoantibody. The two approaches are therefore complementary and needed for future studies

There are clear challenges in dissecting this complex humoral immune response, which comprises both acute and late phases, involves IgM and IgG isotypes that do not always map to expected timelines, and consists of both polyantigenic upregulation and dominant responses to specific antigens (the latter of which involves several neural and non-neural antigens). The polyantigenic IgM upregulation appears to be more easily delineated than dominant responses to specific antigens, and could feasibly be approximated by measuring total serum IgM concentration. If the association with worse outcome is borne out in larger studies, this simple test could represent a useful prognostic biomarker, and perhaps a marker for stratifying patients for immunomodulatory studies.

The additional explanatory power of acute polyantigenic IgM responses in predicting outcome, and the association between anti-MAG IgM and NfL levels months to years after injury, raises the possibility that autoantibodies may be pathogenic. However, direct causality is difficult to confirm, and persistent autoantibody production could also reflect an epiphenomenon caused by ongoing antigen exposure due to neurodegeneration from another mechanism (such as ongoing amyloid or tau-induced neurotoxicity(Johnson *et al*., 2012)). Indeed, these autoantibody responses may even represent an active beneficial response to such ongoing injury. For example, MAG may exert an inhibitory effect on axonal regeneration in adults (McKerracher *et al*., 1994; Mukhopadhyay *et al*., 1994), and rats treated with recombinant anti-MAG monoclonal IgG post-TBI appear to have better outcomes than those treated with mouse IgG (Thompson *et al*., 2006)).

If the autoantibody production is indeed pathogenic, there are several potential routes for such pathogenesis. Firstly, the very presence of large numbers of circulating or deposited immune-complexes can cause tissue injury (Chauhan, 2017). Regarding direct effects, antibodies exert their destructive capabilities through two main mechanisms: complement dependent cytotoxicity and antibody-dependent cellular cytotoxicity (ADCC; the opsonisation and engagement of effector cells). In addition, they can interfere with receptor functioning by blocking ligand-receptor interactions. The levels of intrathecal Membrane Attack Complex (MAC; the key terminal effector of the classical complement pathway) are markedly higher in patients following TBI. Further, a subset of patients display a second peak of intrathecal MAC generation one week after the injury, temporally consistent with the consequence of adaptive immune responses (Stahel *et al*., 2001). Inhibition of the MAC in mice subjected to TBI reduces subsequent secondary neuron loss and promotes neurological recovery (Fluiter *et al*., 2014). While there are no data addressing the effect of TBI-induced autoantibodies on immune cell activation, the related condition of spinal cord injury provides evidence that ADCC is a key driver of secondary injury (Ankeny *et al*., 2009). Given the well-established occurrence of microglial activation following TBI (both acutely and persisting for years post-injury), the interaction between antibody-antigen complexes and the Fcγ receptor on microglia, could represent a bridge between adaptive immune responses and ongoing innate activation (Winter *et al*., 2016).

Therapeutic interventions might be targeted to either prevent the development of autoantibodies early in the disease course with agents such as B-cell activating factor (BAFF)/a proliferation inducing ligand (APRIL) inhibitors or anti-CD20 monoclonal antibodies (Dekaban and Thawer, 2009), or to modulate the long-term persistence of these antibodies with similar medications once the acute phase has passed. Alternatively, if serum or CSF autoantibody levels can identify a subset of patients at risk of complement-mediated injury, they may be used to select patient subsets for targeted trials of complement inhibitors such as eculizumab (Roselli *et al*., 2018).

Further work in this area must focus on two broad questions: 1) what are the implications of autoantibody responses after TBI (elucidated by large observational as well as interventional studies), and 2) what is the underlying biology behind these responses (and hence how can they be best modulated)? Given the heterogeneity of disease and confounding prognostic factors in TBI, large cohorts with detailed clinical information would be necessary to confirm any association between autoantibody status and subsequent outcome, and even then, causality could not be assumed. Transfer of the immunoglobulin fraction from injured animals into either uninjured animals or those prior to experimental TBI would help elucidate the role, although even this would not be able to pick apart the relative effect of the two different responses. Understanding the biology further, particularly that of the concomitant lymphocyte processes would allow for the selective experimental targeting of individual processes. For example, an increase in B1 cells corresponding with polyantigenic responses would support the idea that these do indeed represent “natural” autoantibodies, and similarly B-cells could be challenged with an antigen highlighted by autoantibody screening to clarify antigen-specific responses; each response could then be targeted, and the impact on outcome assessed.

Some limitations of this study should be noted. Firstly, the differences in demographics and outcome data available for the Discovery and Validation cohorts (i.e. different outcome scales used, and timing of follow-up) limit direct comparison, and risk a failure to replicate findings between the cohorts. This may be particularly relevant regarding the comparatively small number of young patients in the Validation cohort (and therefore the number with the greatest magnitude of polyantigenic IgM response) which reduced the power to investigate the association between the polyantigenic IgM response and clinical outcome in this cohort. With regards to the Late and Long-term cohorts, the principal limitation is the use of surrogate biomarkers for neurodegeneration, which, whilst now widely used in the field of neurodegeneration, have not so far been validated in post-TBI neurodegeneration. We did not have separate validation cohorts for the Late and Long-term time-points, and whilst the experimental design protected against batch effects at the point of the microarray assay, we cannot completely exclude effects arising from the initial processing and storage of serum, although this is unlikely to be systematically different between groups. Finally, the protein array platform may not present the target proteins in a natural conformation, and has lower sensitivity than other antibody detection techniques. However, as demonstrated by (i) use of purified immunoglobulin fractionated from sera and (ii) through pre-incubation with cognate antigens, results obtained from the protein microarray were shown to be specific and reproducible.

In conclusion, we have used protein microarray technology to screen for novel autoantibody production following TBI. This approach has elucidated two distinct patterns of response: 1) increase in polyantigenic IgM and IgG, the former of which peaks at day 7, but both of which persist for years post-injury, and 2) dominant autoantibody responses to specific antigens which follow a vaccination-like temporal profile, and persist for months but return to baseline years post-injury. Our data would suggest that the polyantigenic IgM response in the acute phase may be detrimental to clinical outcome, and that persistent anti-MAG IgM autoantibodies associate with surrogate markers of late neurodegeneration.

## Data Availability

All data used in this study are available upon request from the corresponding author.

## Abbreviations

BBB: Blood-brain barrier
CLDN5: Claudin 5
CNS: Central nervous system
COL5a2: Collagen type 5 alpha 2 chain
DAMP: Danger-associated molecular pattern
GFAP: Glial fibrillary acidic protein
GOS: Glasgow Outcome Score
GOSE: Glasgow Outcome Score Extended
Ig: Immunoglobulin
MAC: Membrane attack complex
MAG: Myelin-associated glycopeptide
MBP: Myelin basic protein
MFI: Median fluorescence intensity
NfL: Neurofilament light
SCI: Spinal cord injury
SELE: E-selectin
TBI: Traumatic brain injury
TJP-1: Tight junction protein 1

## Acknowledgements

We thank Paddy Waters (Neuroimmunology Group, Nuffield Department of Clinical Neurosciences, John Radcliffe Hospital, Oxford, UK), and Sanja Ugrinovic (Department of Infectious Diseases, Cambridge University Hospitals, Cambridge, UK) for sharing samples which contributed to the development of the protein microarray and normative data. We gratefully acknowledge the participation of all NIHR Cambridge BioResource volunteers, and thank the NIHR Cambridge BioResource centre and staff for their contribution. We thank the National Institute for Health Research and NHS Blood and Transplant.

## Funding

EJN, AH, AJC, PJH and DKM are supported by the Medical Research Council (UK) within the framework of ERA-NET NEURON. ERZ is supported by the Ministero della Salute (Italy) within the framework of ERA-NET NEURON. DKM is supported by an NIHR Senior Investigator Award and European Union 7th Framework program (EC grant <u>602150</u>). EJN is supported by the *Intensive Care Society Young Investigator Award, Addenbrookes Charitable Trust Cambridge Clinical Research Fellowship and NIHR* Brain Injury Healthcare Technology Co-operative Seedcorn funding. EPT acknowledges funding from Region Stockholm (ALF), the Swedish Brain Foundation (Hjärnfonden) and the Swedish Society for Medical Research (Svenska Sällskapet för Medicinsk Forskning). HZ is a Wallenberg Scholar supported by grants from the Swedish Research Council (#2018-02532), the European Research Council (#681712), Swedish State Support for Clinical Research (#ALFGBG-720931), Centrum för Idrottsforskning (#P2019-0198), and the UK Dementia Research Institute at UCL. PJH is funded by the NIHR (Research Professorship and Cambridge BRC) and Royal College of Surgeons of England. JJ is funded by the Wellcome Trust (RG79413). VFJN is funded by an Academy of Medical Sciences / The Health Foundation Clinician Scientist Fellowship. HSM is supported by the Cambridge Trust and the Rosetrees Trust

## Competing Interests

HZ has served at scientific advisory boards for Denali, Roche Diagnostics, Wave, Samumed and CogRx, has given lectures in symposia sponsored by Fujirebio, Alzecure and Biogen, and is a co-founder of Brain Biomarker Solutions in Gothenburg AB, a GU Ventures-based platform company at the University of Gothenburg. AJC received honoraria and travel expenses from Genzyme [a Sanofi company] until September 2017. VFJN reports personal fees from Neurodiem, outside the submitted work.

DKM reports grants, personal fees, and non-financial support from GlaxoSmithKline Ltd., grants, personal fees, and other from NeuroTrauma Sciences, grants and personal fees from Integra Life Sciences, personal fees from Pfizer Ltd., grants and personal fees from Lantmannen AB, from Calico Ltd., personal fees from Pressura Neuro Ltd., others from Cortirio Ltd., outside the submitted work. MT is the founder and CEO of Cambridge Protein Arrays Ltd., a company which performs antibody specificity profiling.

## Supplementary Figures

**Supplementary Figure 1.**
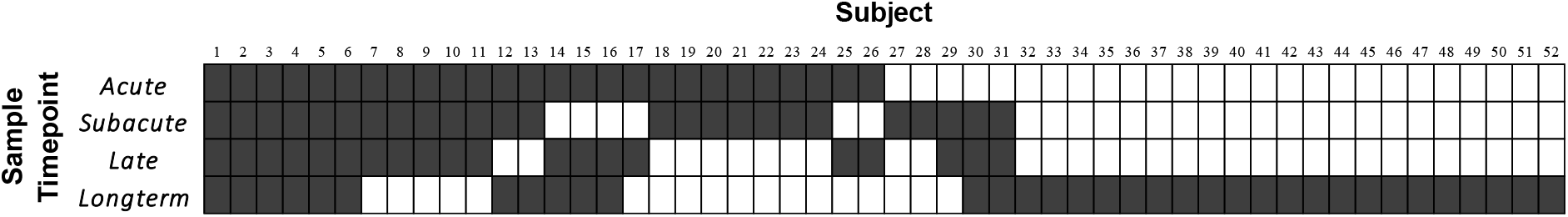
Schematic diagram displaying how patients from the Discovery cohort contributed to the Late and Long-term cohorts. *Shaded boxes represent time-points where samples were taken from individuals*.

**Supplementary Figure 2.**
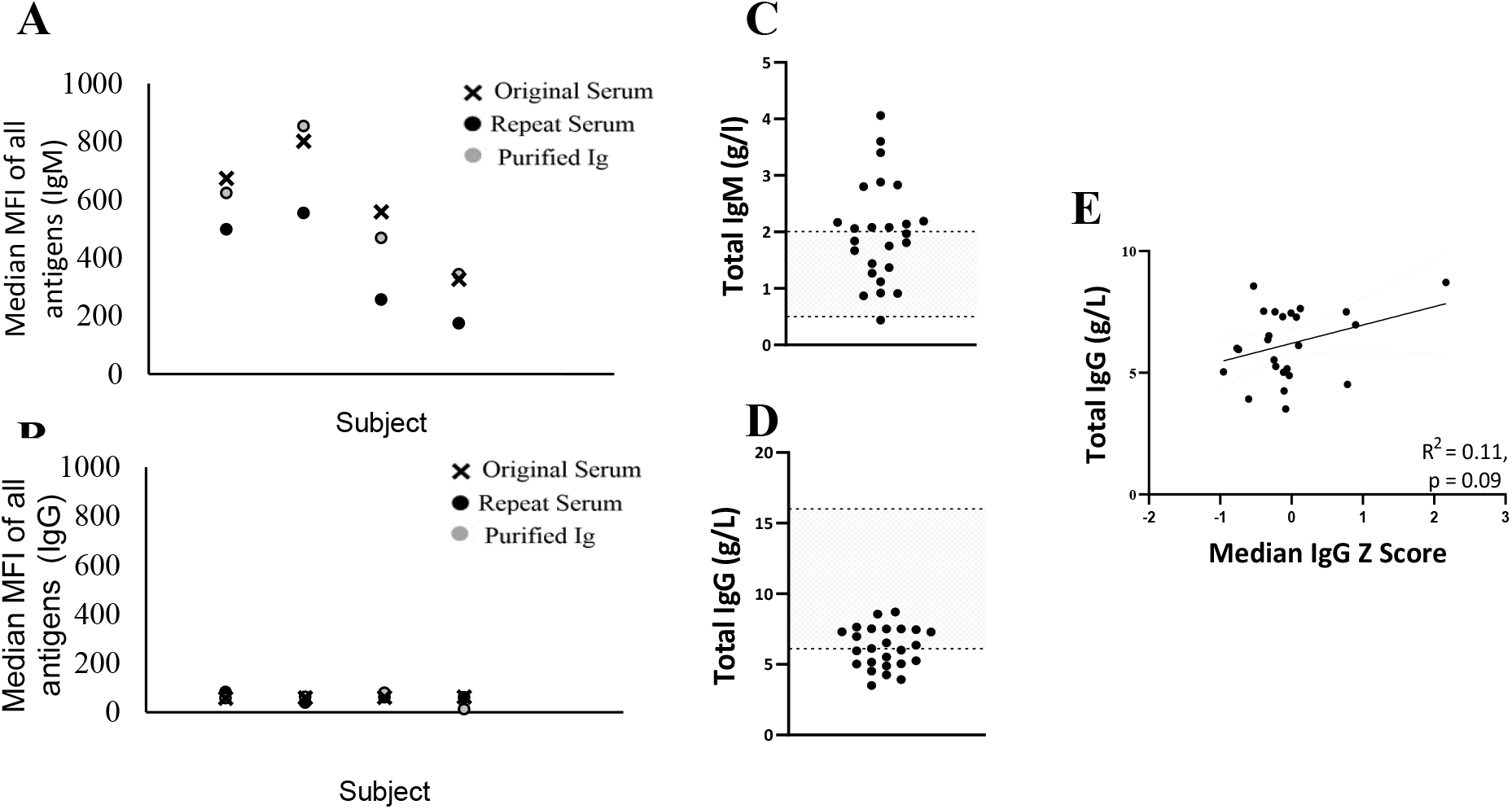
(**A**) Polyantigenic IgM response is replicated both in a technical repeat (serum assayed again in a separate experiment) and in the purified immunoglobulin fraction (**B**) no such replication is seen with IgG (**C**) Total serum IgM concentrations was above the normal range in 12/25 patients. *Hatched area = normal range* (**D**) Total serum IgG concentration was below the normal range in 13/25 patients. *Hatched area = normal range* (**E**) Median IgG Z score did not correlate with total serum IgG

**Supplementary Figure 3.**
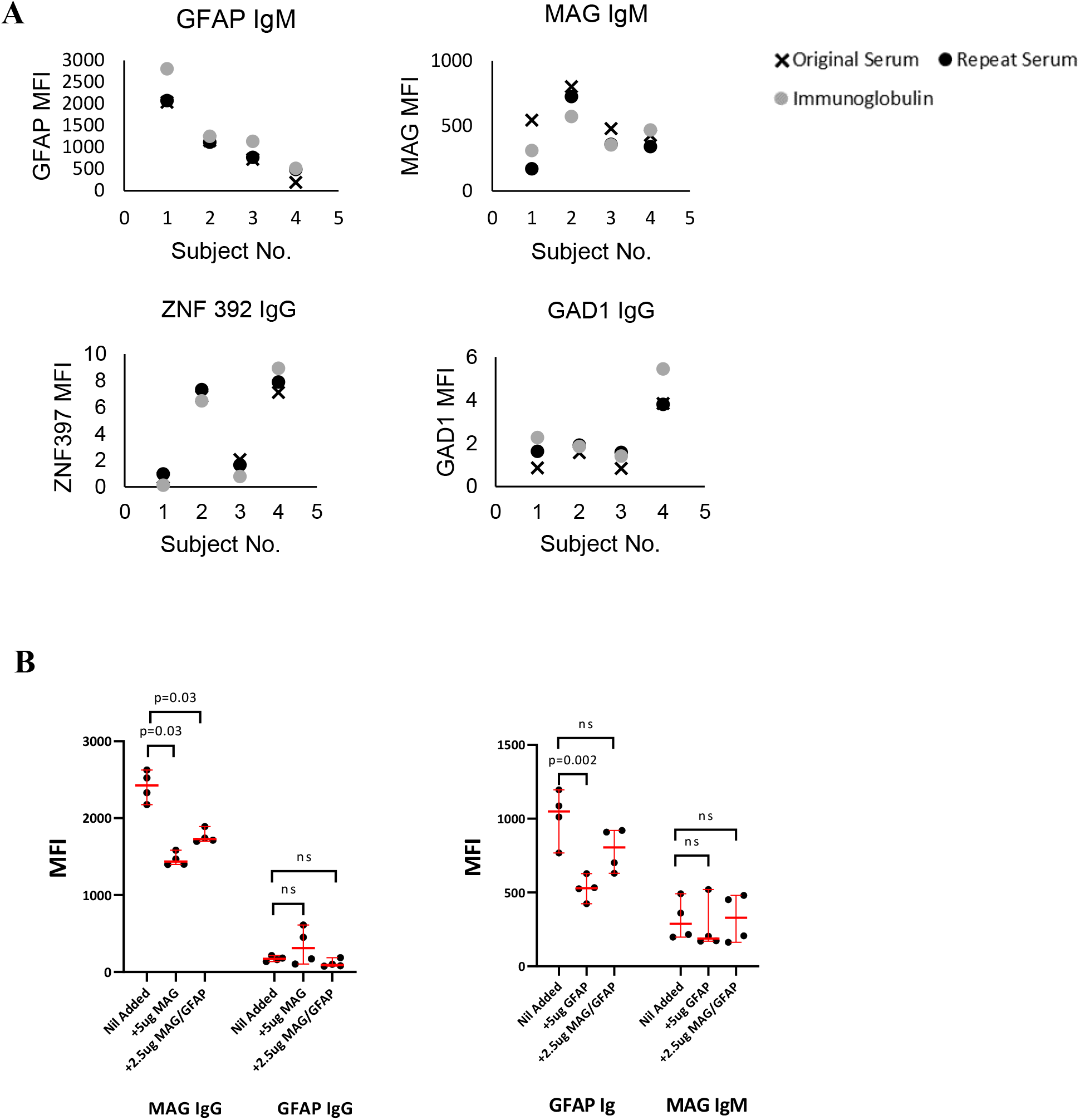
(**A**) Graphs displaying replication of dominant responses to specific antigens both in a technical repeat (serum assayed again in a separate experiment) and in the purified immunoglobulin fraction (**B**) Graphs displaying the effect on MFI when serum was pre-incubated with the cognate antigen. The sample positive for anti-MAG IgG antibodies showed a reduction in MFI when preincubated with MAG protein; the corresponding GFAP IgG MFI in the same sample did not reduce on pre-incubation reassuring against a sump effect from adding protein in general. The sample positive for anti-GFAP IgM showed a similar reduction in MFI when pre-incubated with GFAP protein; again, the corresponding MAG MFI did not reduce when pre-incubated with GFAP protein.

**Supplementary Figure 4.**
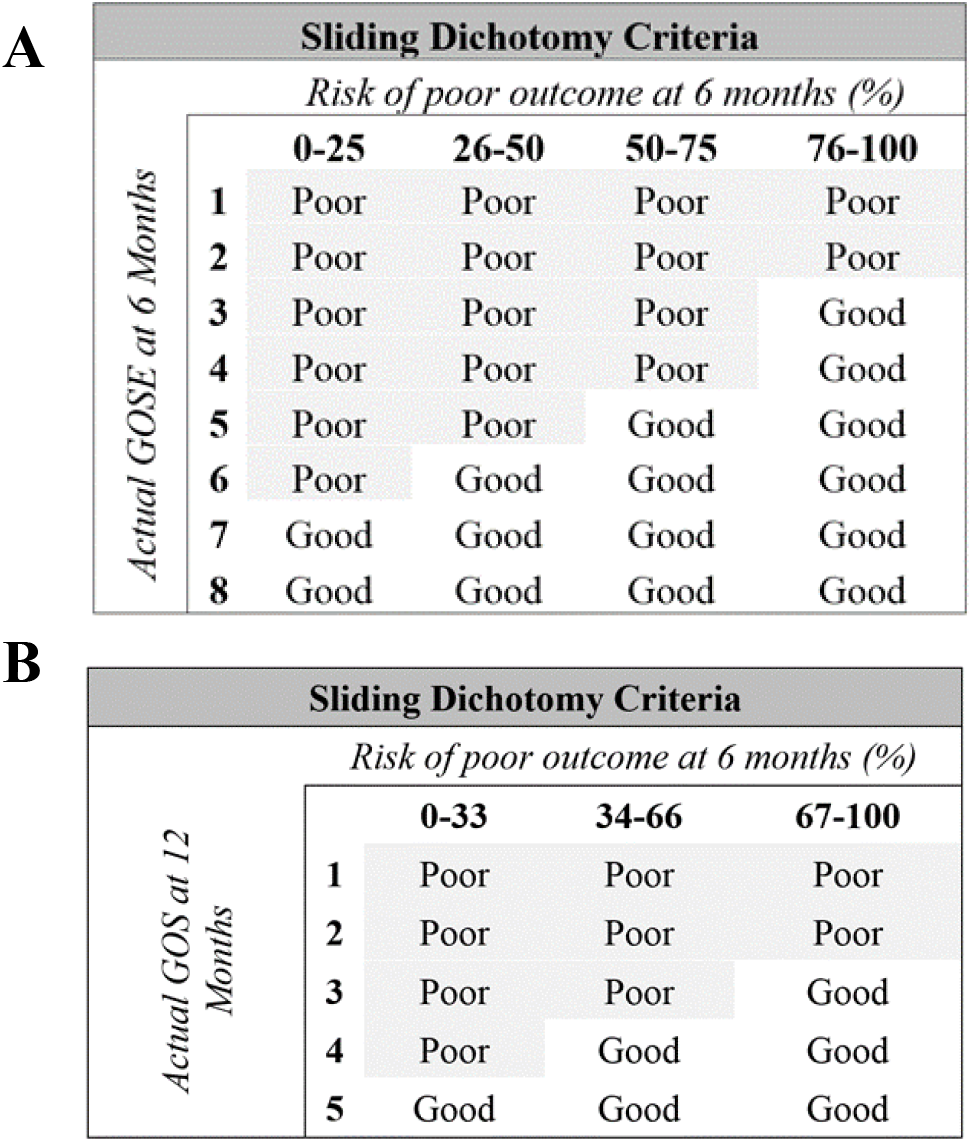
Criteria used to dichotomise patients’ outcome into “worse than expected” and “as/better than expected” groups, according to their risk of poor outcome as judged by the IMPACT score variables for the (**A**) Discovery cohort and (**B**) Validation cohort

**Supplementary Figure 5.**
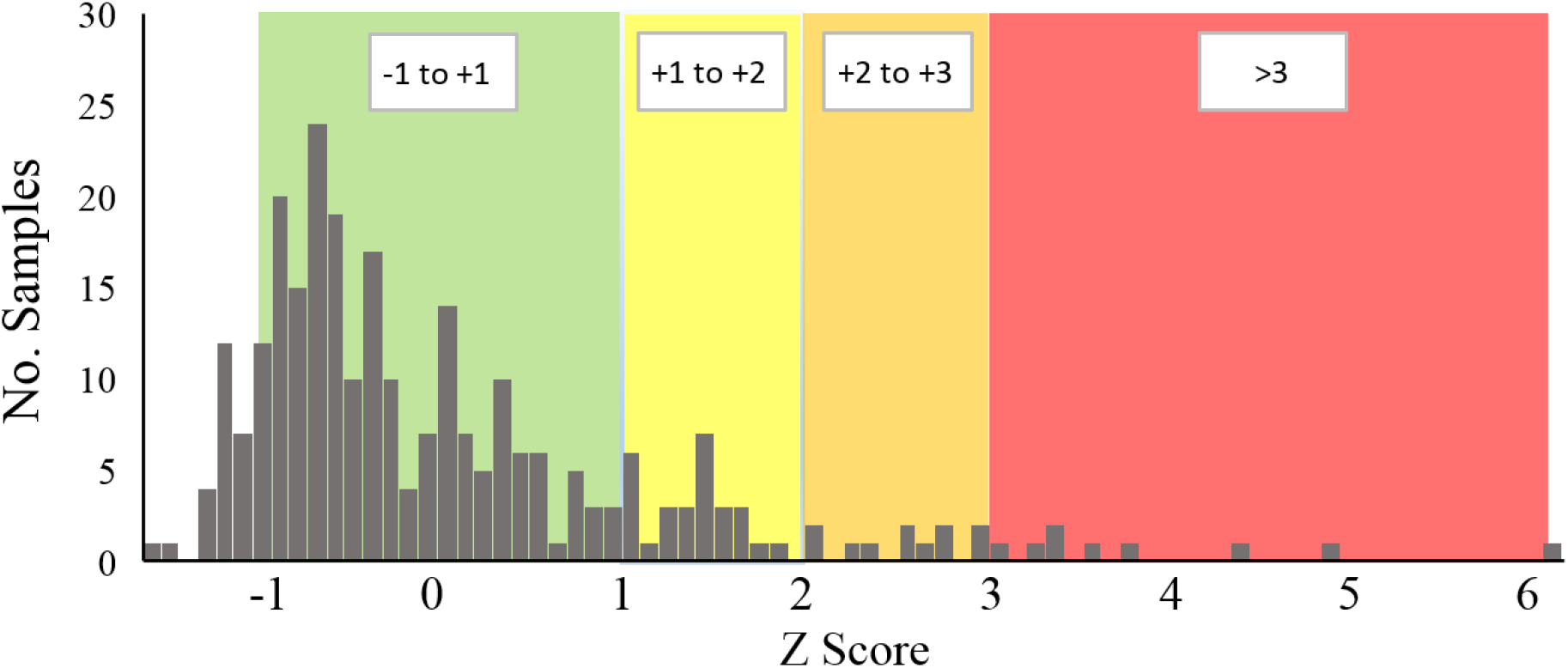
Histogram displaying the distribution of Z scores of IgG MAG MFI in all samples used to generate the reference distributions.

**Supplementary Table 1.**
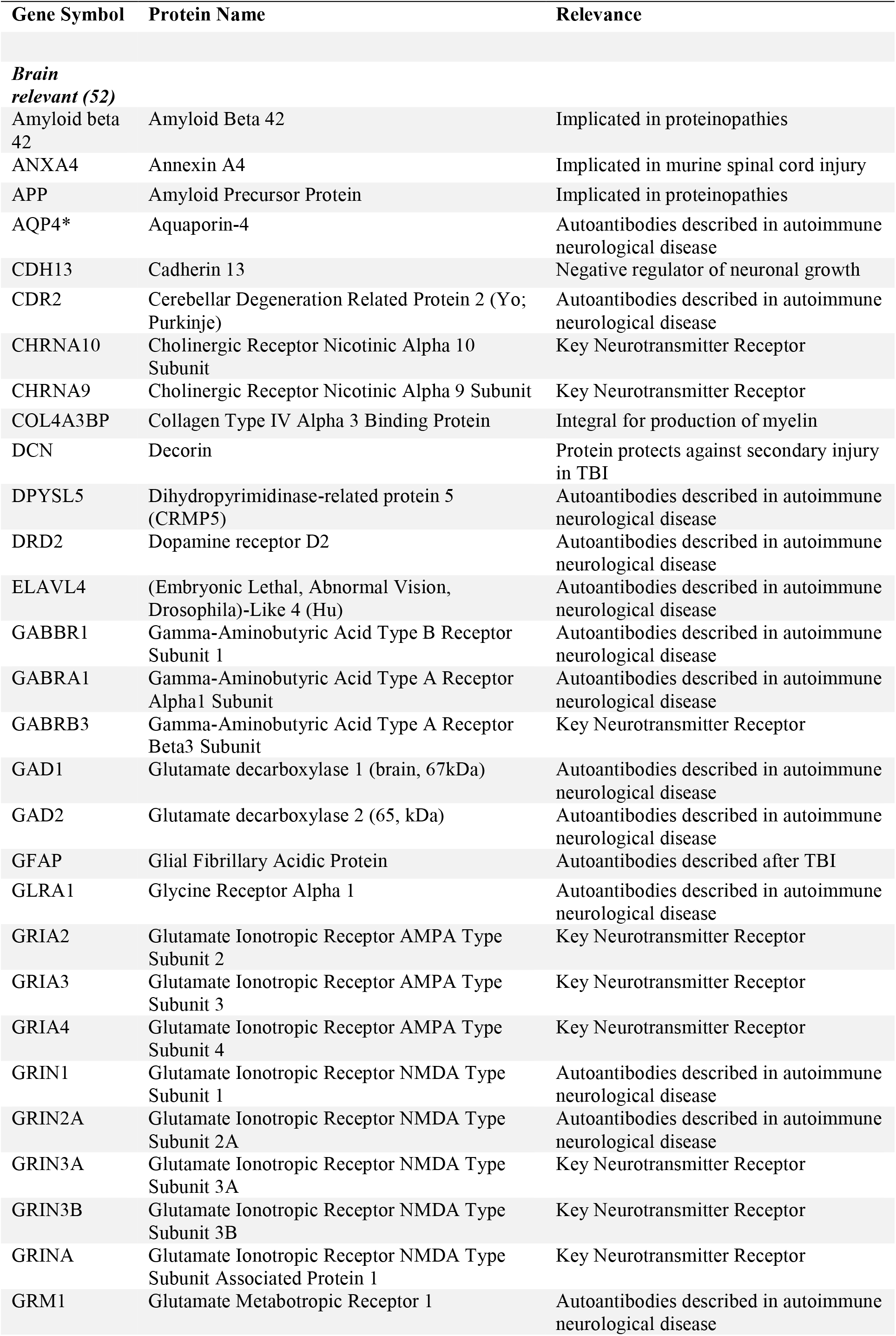
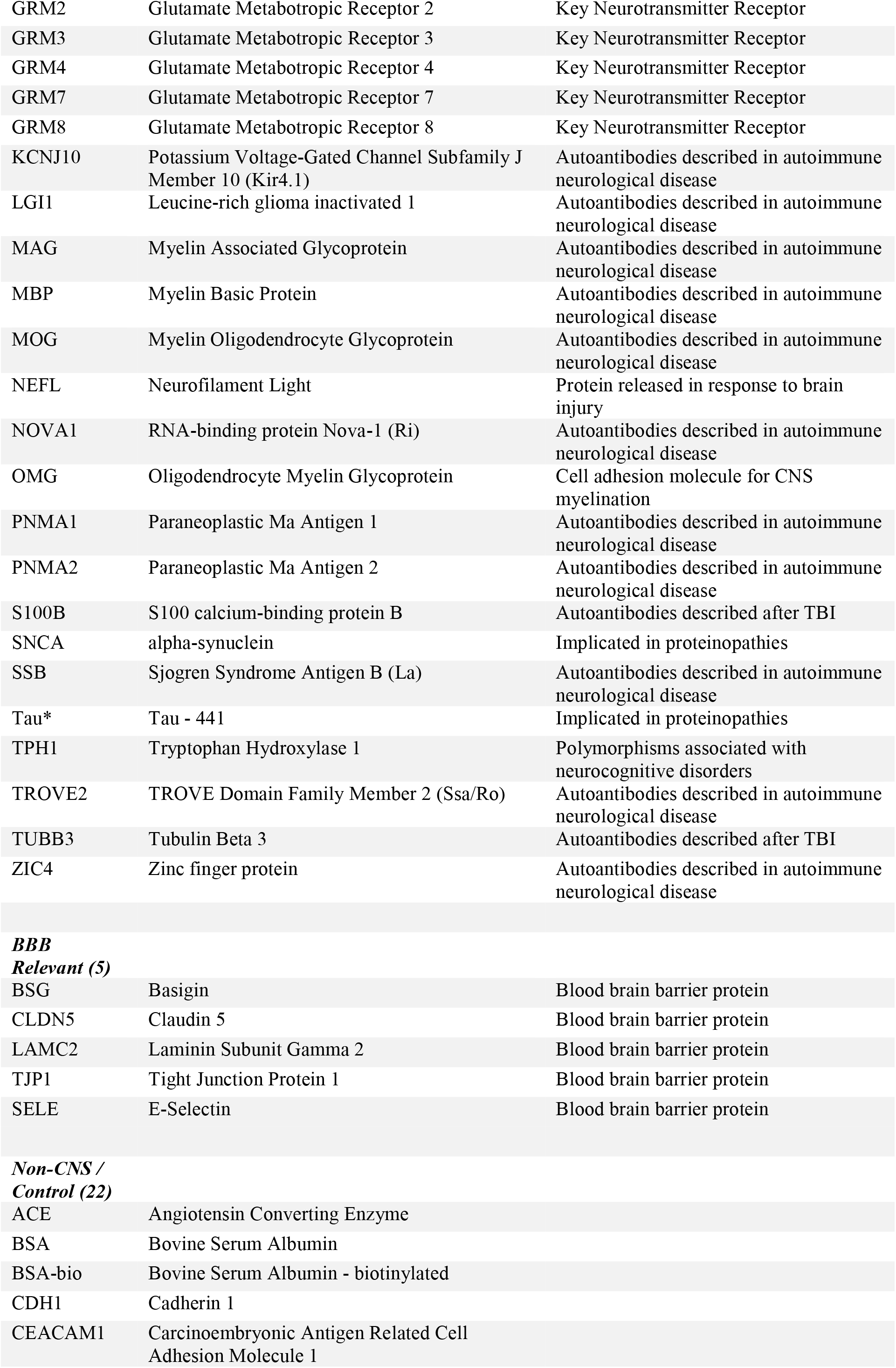
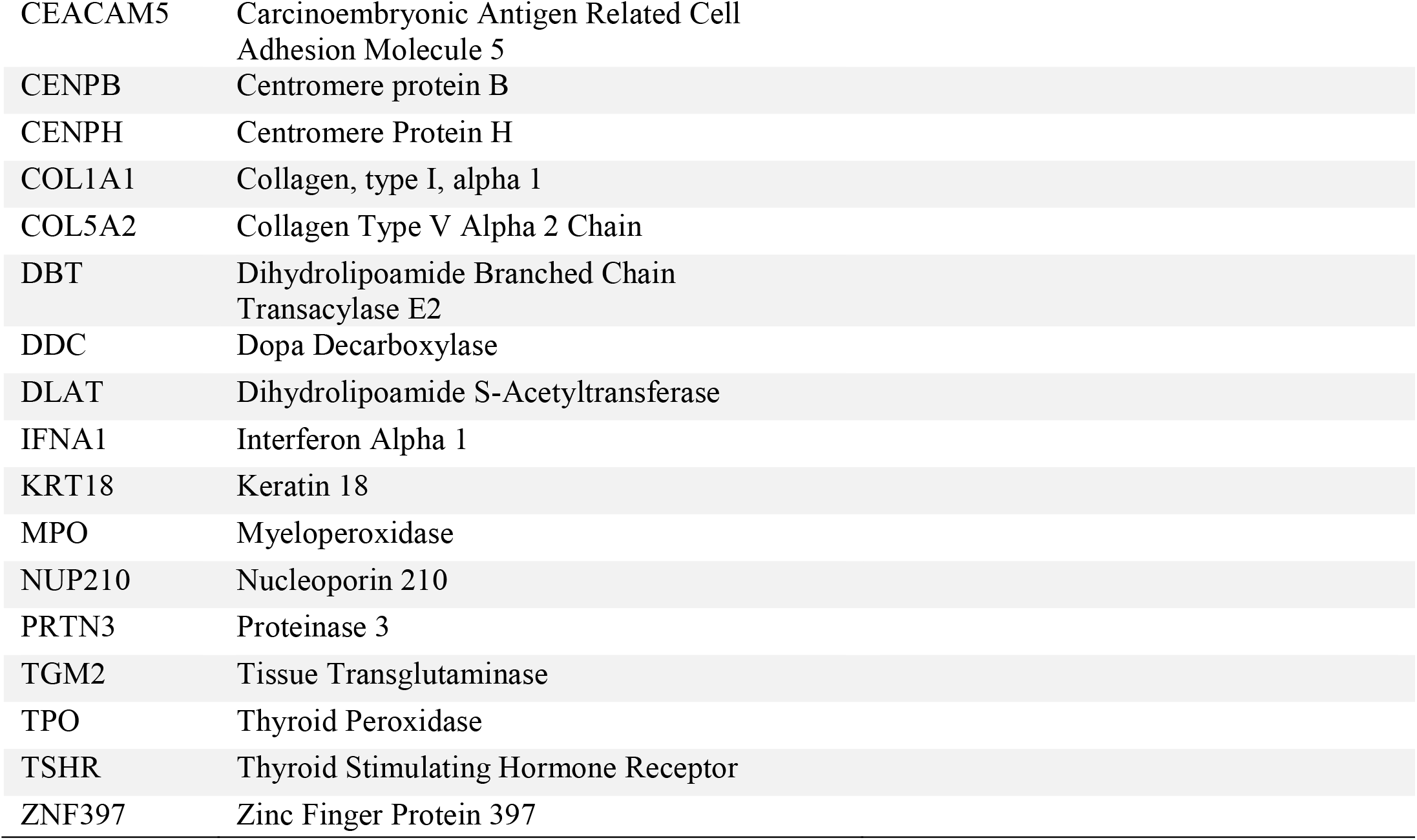
Custom Protein Microarray Protein List. Full list of antigens printed on custom central nervous system protein microarray

**Supplementary Table 2.**
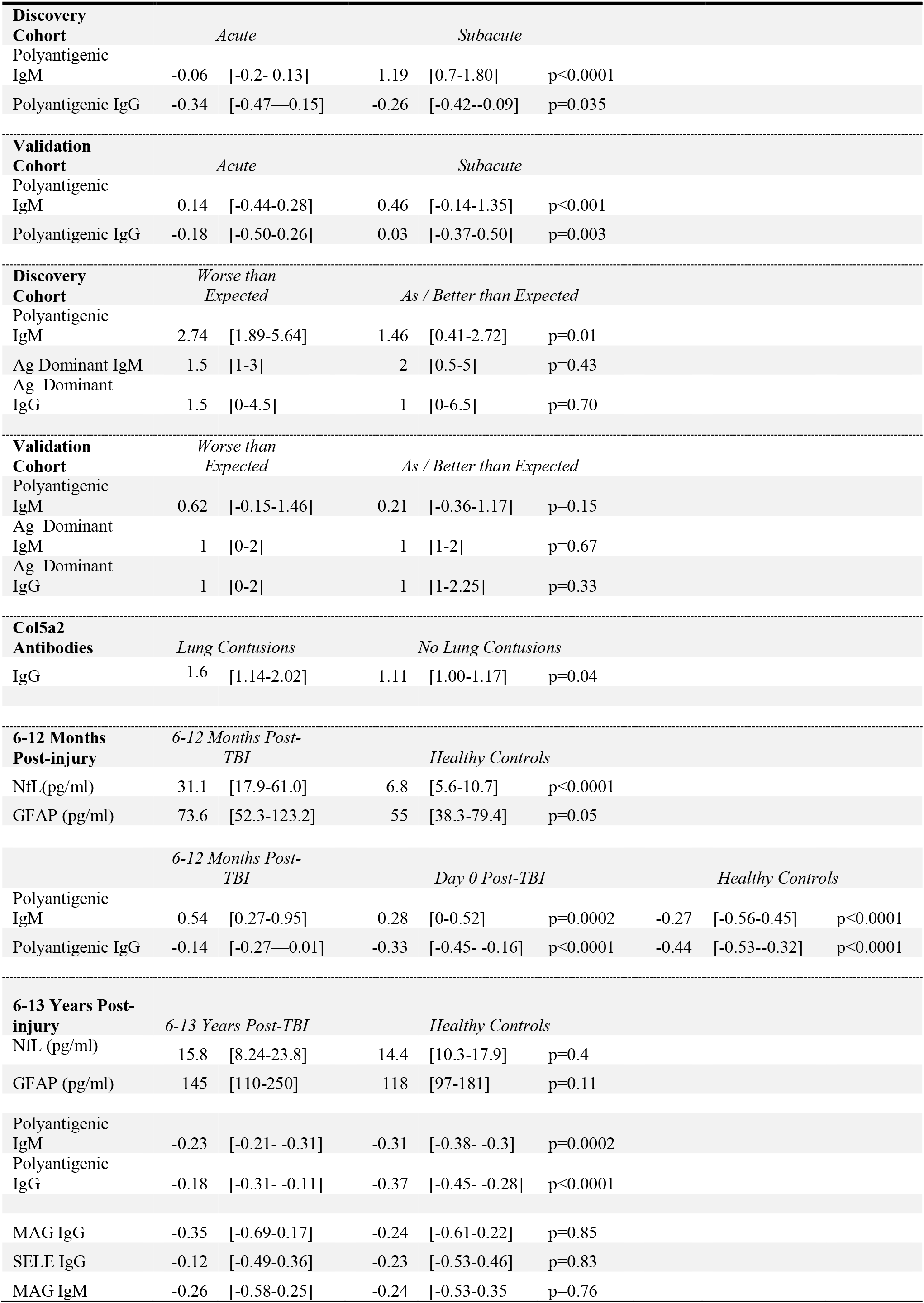
Statistics for All Group Comparisons. Values for statistics quoted in body of main text. “Polyantigenic” values refer to the median Z score per patient, “Ag (antigen) dominant” values refers to the number of dominant antibodies against specific antigens detected per patient. MAG/SELE values refer to the Z scores of the particular antigen-specific autoantibody. All values represent Median [IQR].

**Supplementary Table 3.** Comparison of whole group Z score change between Acute and Subacute samples for the 5 most frequently seen autoantibodies (**A&B**), and again but with Z>3 results removed still showing differences between (**C&D**) * denotes p-values which remain significant following Benjamini-Hochberg correction.

**A**
| Autoantibody Target (IgG) | Acute Median Z-Score (IQR) | Subacute Median Z-Score (IQR) | P-value |
| --- | --- | --- | --- |
| CLDN5 | -0.24<br>(-0.73-0.39) | -0.19<br>(-0.55-0.44) | 0.18 |
| COL5a2 | -0.27<br>(-0.73-0.18) | -0.33<br>(-0.80-0.25) | 0.71 |
| GFAP | -0.36<br>(-0.69-0.15) | -0.15<br>(-0.71-0.46) | 0.19 |
| MAG | -0.16<br>(-0.75-0.28) | 0.02<br>(-0.53-0.62) | **0.001\*** |
| SELE | -0.29<br>(-0.65-0.35) | -0.28<br>(-0.64-0.36) | 0.41 |

**B**
| Autoantibody Target (IgM) | Acute Median Z-Score (IQR) | Subacute Median Z-Score (IQR) | P-value |
| --- | --- | --- | --- |
| CLDN5 | 0.45<br>(-0.14-1.24) | 0.30<br>(-0.54-0.97) | 0.12 |
| COL5a2 | 0.07<br>(-0.48-0.56) | 0.25<br>(-0.40-1.00) | **0.005\*** |
| GFAP | -0.13<br>(-0.78-0.72) | -0.13<br>(-0.73-1.25) | **0.04\*** |
| MAG | 0.05<br>(-0.70-0.80) | 0.61<br>(-0.13-1.22) | **0.004\*** |
| SELE | 0.26<br>(-0.50-0.89) | 0.16<br>(-0.59-1.10) | 0.22 |

**C**
| Autoantibody Target (IgG) | Acute Median Z-Score (IQR) | Subacute Median Z-Score (IQR) | P-value |
| --- | --- | --- | --- |
| CLDN5 | -0.19<br>(-0.71-0.47) | -0.16<br>(-0.54-0.60) | 0.13 |
| COL5a2 | -0.24<br>(-0.73-0.20) | -0.30<br>(-0.79-0.48) | 0.40 |
| GFAP | -0.33<br>(-0.67-0.22) | -0.12<br>(-0.69-0.61) | 0.18 |
| MAG | -0.14<br>(-0.69-0.33) | -0.05<br>(-0.52-0.77) | **0.0007\*** |
| SELE | -0.27<br>(-0.65-0.43) | -0.17<br>(-0.63-0.61) | 0.1 |

| Autoantibody Target (IgM) | Acute Median Z-Score (IQR) | Subacute Median Z-Score (IQR) | P-value |
| --- | --- | --- | --- |
| CLDN5 | 0.45<br>(-0.14-1.24) | 0.36<br>(-0.51-1.06) | 0.36 |
| COL5a2 | 0.19<br>(-0.44-0.59) | 0.33<br>(-0.35-1.19) | **0.0007\*** |
| GFAP | -0.1<br>(-0.77-0.77) | -0.1<br>(0.70-1.38) | **0.0168** |
| MAG | 0.3 (-0.48-0.93) | 0.6 (-0.38-2.09) | **0.008\*** |
| SELE | 0.05<br>(-0.69-0.83) | 0.65<br>(-0.002-1.3) | **0.0018\*** |

**Supplementary Table 4.** Characteristics of Chronic TBI and Control Cohorts. Baseline characteristics of Late (6-12 month post-injury) and Long-term (6-13 years post-injury) groups and their respective healthy control groups. P-value (a) refers to the comparison between the Late TBI cohort and Control Group 1, and p-value (b) to the comparison between the Long-term TBI cohort and Control Group 2.

| Characteristic | Late TBI Cohort (n=20) | Control Group 1 (n=17) | p-value (a) | Long-term TBI (n=34) | Control Group 2 (n=28) | p-value (b) |
| --- | --- | --- | --- | --- | --- | --- |
| Sex - No. Male | 18 (90) | 6 (35) | 0.001 | 23 (65) | 20 (71) | 0.79 |
| Age (Yr) | 30 (25-55) | 39 (28-42) | 0.1 | 52 (31-65) | 50 (38-57) | 0.53 |

**Supplementary Table 5.** Screening for the 5 most frequently seen autoantibodies in the acute phase reveals persistent IgM to MAG at a group level. * denotes p-values which remain significant following the Benjamini-Hochberg procedure.

| Autoantibody Target (IgM) | Healthy Control | Acute | Late | Healthy Control vs. Late p-value | Early vs. Late post-TBI p-value |
| --- | --- | --- | --- | --- | --- |
| CLDN5 | -0.76<br>(-1.40—0.08) | -0.19<br>(-1.12-0.53) | -0.77<br>(1.18-0.09) | 0.50 | 0.92 |
| COL5a2 | -0.06<br>(-0.87-0.69) | -0.05<br>(-0.97-0.28) | -0.14<br>(-0.86-0.92) | 0.92 | 0.96 |
| GFAP | 0.28<br>(-0.11-0.97) | 0.35<br>(-0.26-0.90) | 0.71<br>(-0.23-1.44) | 0.17 | 0.27 |
| MAG | -0.29<br>(-0.84-0.47) | -0.26<br>(-0.88-0.82) | 0.43<br>(-0.4-1.63) | 0.03 | **0.008\*** |
| SELE | 0.06<br>(-0.61-0.89) | 0.04<br>(-0.23-0.74) | 0<br>(-0.32-0.76) | 0.97 | 0.85 |

**Supplementary Table 6.**
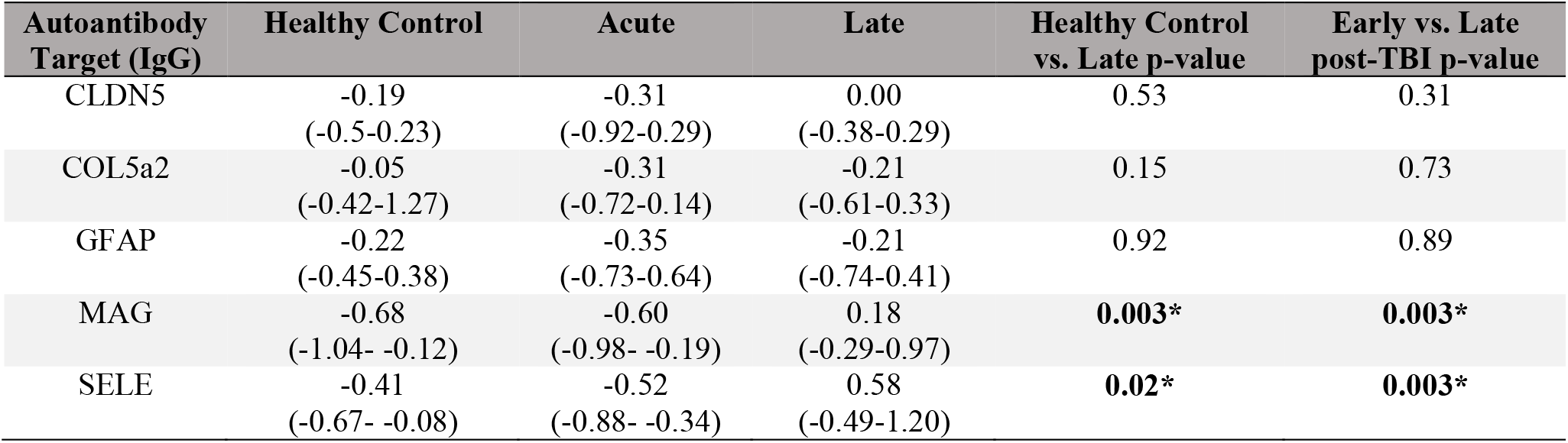
Screening for the 5 most frequently seen autoantibodies in the acute phase reveals persistent IgG to MAG and SELE at a group level. * denotes p-values which remain significant following the Benjamini-Hochberg procedure.

## Notes

### Author Declarations

The studies described were approved either by the Cambridgeshire local research ethics committee (REC 97/290 and 13/EE/0119), or regional ethical board in Stockholm (#2005/1526/31/2).

## References

Absinta M, Ha S-K, Nair G, Sati P, Luciano NJ, Palisoc M, et al. Human and nonhuman primate meninges harbor lymphatic vessels that can be visualized noninvasively by MRI [Internet]. eLife 2017; 6[cited 2018 Jan 24] Available from: https://www.ncbi.nlm.nih.gov/pmc/articles/PMC5626482/

Ankeny DP, Guan Z, Popovich PG. B cells produce pathogenic antibodies and impair recovery after spinal cord injury in mice. J Clin Invest 2009; 119: 2990–9.

Ankeny DP, Lucin KM, Sanders VM, McGaughy VM, Popovich PG. Spinal cord injury triggers systemic autoimmunity: evidence for chronic B lymphocyte activation and lupus-like autoantibody synthesis. J Neurochem 2006; 99: 1073–87.

Baker SP, O’Neill B, Haddon W, Long WB. The injury severity score: a method for describing patients with multiple injuries and evaluating emergency care. J Trauma 1974; 14: 187–96.

Bazarian JJ, Biberthaler P, Welch RD, Lewis LM, Barzo P, Bogner-Flatz V, et al. Serum GFAP and UCH-L1 for prediction of absence of intracranial injuries on head CT (ALERT-TBI): a multicentre observational study. Lancet Neurol 2018; 17: 782–9.

Chauhan AK. Editorial: Immune Complexes in Disease Pathology [Internet]. Front Immunol 2017; 8[cited 2020 Mar 24] Available from: https://www.frontiersin.org/articles/10.3389/fimmu.2017.00173/full

Cox AL, Coles AJ, Nortje J, Bradley PG, Chatfield DA, Thompson SJ, et al. An investigation of auto- reactivity after head injury. J Neuroimmunol 2006; 174: 180–6.

Dekaban GA, Thawer S. Pathogenic antibodies are active participants in spinal cord injury. J Clin Invest 2009; 119: 2881–4.

Dodel R, Balakrishnan K, Keyvani K, Deuster O, Neff F, Andrei-Selmer L-C, et al. Naturally Occurring Autoantibodies against β-Amyloid: Investigating Their Role in Transgenic Animal and In Vitro Models of Alzheimer’s Disease. J Neurosci 2011; 31: 5847–54.

Fluiter K, Opperhuizen AL, Morgan BP, Baas F, Ramaglia V. Inhibition of the Membrane Attack Complex of the Complement System Reduces Secondary Neuroaxonal Loss and Promotes Neurologic Recovery after Traumatic Brain Injury in Mice. J Immunol 2014; 192: 2339–48.

Gelfand JM. Autoimmune encephalitis after herpes simplex encephalitis: insights into pathogenesis. Lancet Neurol 2018; 17: 733–5.

Greenberg G, Mikulis DJ, Ng K, DeSouza D, Green RE. Use of Diffusion Tensor Imaging to Examine Subacute White Matter Injury Progression in Moderate to Severe Traumatic Brain Injury. Arch Phys Med Rehabil 2008; 89: S45–50.

Grönwall C, Vas J, Silverman GJ. Protective Roles of Natural IgM Antibodies [Internet]. Front Immunol 2012; 3[cited 2019 Oct 14] Available from: https://www.frontiersin.org/articles/10.3389/fimmu.2012.00066/full

Güven E, Duus K, Lydolph MC, Jørgensen CS, Laursen I, Houen G. Non-specific binding in solid phase immunoassays for autoantibodies correlates with inflammation markers. J Immunol Methods 2014; 403: 26–36.

Hammond FM, Grattan KD, Sasser H, Corrigan JD, Rosenthal M, Bushnik T, et al. Five years after traumatic brain injury: A study of individual outcomes and predictors of change in function. NeuroRehabilitation 2004; 19: 25–35.

Himanen L, Portin R, Isoniemi H, Helenius H, Kurki T, Tenovuo O. Longitudinal cognitive changes in traumatic brain injury: a 30-year follow-up study. Neurology 2006; 66: 187–92.

Iwata T, Philipovskiy A, Fisher AJ, Presson RG, Chiyo M, Lee J, et al. Anti-Type V Collagen Humoral Immunity in Lung Transplant Primary Graft Dysfunction. J Immunol Baltim Md 1950 2008; 181: 5738–47.

Jennett B. Epidemiology of head injury. J Neurol Neurosurg Psychiatry 1996; 60: 362–369.

Jennett B, Bond M. ASSESSMENT OF OUTCOME AFTER SEVERE BRAIN DAMAGE: A Practical Scale. The Lancet 1975; 305: 480–4.

Jeong JS, Jiang L, Albino E, Marrero J, Rho HS, Hu J, et al. Rapid Identification of Monospecific Monoclonal Antibodies Using a Human Proteome Microarray [Internet]. Mol Cell Proteomics MCP 2012; 11[cited 2020 Jan 8] Available from: https://www.ncbi.nlm.nih.gov/pmc/articles/PMC3433917/

Johnson VE, Stewart JE, Begbie FD, Trojanowski JQ, Smith DH, Stewart W. Inflammation and white matter degeneration persist for years after a single traumatic brain injury. Brain 2013; 136: 28–42.

Johnson VE, Stewart W, Smith DH. Widespread Tau and Amyloid-Beta Pathology Many Years After a Single Traumatic Brain Injury in Humans. Brain Pathol 2012; 22: 142–9.

Jr ETC, Kilmartin D, Agarwal M, Zierhut M. Sympathetic Ophthalmia. Ocul Immunol Inflamm 2017; 25: 149–51.

Kumar R, Husain M, Gupta RK, Hasan KM, Haris M, Agarwal AK, et al. Serial Changes in the White Matter Diffusion Tensor Imaging Metrics in Moderate Traumatic Brain Injury and Correlation with Neuro-Cognitive Function. J Neurotrauma 2009; 26: 481–95.

Lingsma HF, Roozenbeek B, Steyerberg EW, Murray GD, Maas AI. Early prognosis in traumatic brain injury: from prophecies to predictions. Lancet Neurol 2010; 9: 543–54.

Maas AI, Murray G, Henney H, Kassem N, Legrand V, Mangelus M, et al. Efficacy and safety of dexanabinol in severe traumatic brain injury: results of a phase III randomised, placebo-controlled, clinical trial. Lancet Neurol 2006; 5: 38–45.

Marchi N, Bazarian JJ, Puvenna V, Janigro M, Ghosh C, Zhong J, et al. Consequences of Repeated Blood-Brain Barrier Disruption in Football Players. PLoS ONE 2013; 8: e56805.

Marshall LF, Marshall SB, Klauber MR, Van Berkum Clark M, Eisenberg H, Jane JA, et al. The diagnosis of head injury requires a classification based on computed axial tomography. J Neurotrauma 1992; 9 Suppl 1: S287–292.

McKerracher L, David S, Jackson DL, Kottis V, Dunn RJ, Braun PE. Identification of myelin-associated glycoprotein as a major myelin-derived inhibitor of neurite growth. Neuron 1994; 13: 805–11.

Millis SR, Rosenthal M, Novack TA, Sherer M, Nick TG, Kreutzer JS, et al. Long-term neuropsychological outcome after traumatic brain injury. J Head Trauma Rehabil 2001; 16: 343–355.

Mukhopadhyay G, Doherty P, Walsh FS, Crocker PR, Filbin MT. A novel role for myelin-associated glycoprotein as an inhibitor of axonal regeneration. Neuron 1994; 13: 757–67.

Murray GD, Barer D, Choi S, Fernandes H, Gregson B, Lees KR, et al. Design and Analysis of Phase III Trials with Ordered Outcome Scales: The Concept of the Sliding Dichotomy. J Neurotrauma 2005; 22: 511–7.

Newcombe VFJ, Correia MM, Ledig C, Abate MG, Outtrim JG, Chatfield D, et al. Dynamic Changes in White Matter Abnormalities Correlate With Late Improvement and Deterioration Following TBI: A Diffusion Tensor Imaging Study. Neurorehabil Neural Repair 2016; 30: 49–62.

Ngankam L, Kazantseva NV, Gerasimova MM. [Immunological markers of severity and outcome of traumatic brain injury]. Zhurnal Nevrol Psikhiatrii Im SS Korsakova Minist Zdr Meditsinskoĭ Promyshlennosti Ross Fed Vserossiĭskoe Obshchestvo Nevrol Vserossiĭskoe Obshchestvo Psikhiatrov 2011; 111: 61–5.

Palma J, Tokarz-Deptuła B, Deptuła J, Deptuła W. Natural antibodies – facts known and unknown. Cent-Eur J Immunol 2018; 43: 466–75.

Panda S, Zhang J, Tan NS, Ho B, Ding JL. Natural IgG antibodies provide innate protection against ficolin-opsonized bacteria. EMBO J 2013; 32: 2905–19.

Plog BA, Dashnaw ML, Hitomi E, Peng W, Liao Y, Lou N, et al. Biomarkers of Traumatic Injury Are Transported from Brain to Blood via the Glymphatic System. J Neurosci 2015; 35: 518–26.

Ramlackhansingh AF, Brooks DJ, Greenwood RJ, Bose SK, Turkheimer FE, Kinnunen KM, et al. Inflammation after trauma: Microglial activation and traumatic brain injury. Ann Neurol 2011; 70: 374–83.

Rodriguez-Zhurbenko N, Quach TD, Hopkins TJ, Rothstein TL, Hernandez AM. Human B-1 Cells and B-1 Cell Antibodies Change With Advancing Age [Internet]. Front Immunol 2019; 10[cited 2020 Jan 13] Available from: https://www.frontiersin.org/articles/10.3389/fimmu.2019.00483/full

Roselli F, Karasu E, Volpe C, Huber-Lang M. Medusa’s Head: The Complement System in Traumatic Brain and Spinal Cord Injury. J Neurotrauma 2018; 35: 226–40.

Ruff RM, Young D, Gautille T, Marshall LF, Barth J, Jane JA, et al. Verbal learning deficits following severe head injury: heterogeneity in recovery over 1 year. Spec Suppl 1991; 75: S50–8.

Sander AM, Roebuck TM, Struchen MA, Sherer M, High Jr WM. Long-Term Maintenance of Gains Obtained in Postacute Rehabilitation by Persons with Traumatic Brain Injury. J Head Trauma Rehabil 2001; 16: 356–373.

Sasaki S, Sullivan M, Narvaez CF, Holmes TH, Furman D, Zheng N-Y, et al. Limited efficacy of inactivated influenza vaccine in elderly individuals is associated with decreased production of vaccine-specific antibodies. J Clin Invest 2011; 121: 3109–19.

Scott G, Hellyer PJ, Ramlackhansingh AF, Brooks DJ, Matthews PM, Sharp DJ. Thalamic inflammation after brain trauma is associated with thalamo-cortical white matter damage [Internet]. J Neuroinflammation 2015; 12[cited 2016 Sep 29] Available from: http://www.ncbi.nlm.nih.gov/pmc/articles/PMC4666189/

Scott G, Zetterberg H, Jolly A, Cole JH, De Simoni S, Jenkins PO, et al. Minocycline reduces chronic microglial activation after brain trauma but increases neurodegeneration. Brain 2018; 141: 459–71.

Shamreĭ RK. [The value of determining autoantibodies in the diagnosis and expertise of closed brain injury]. Voen-Medit?s?inskiĭ Zhurnal 1969; 4: 39–43.

Škoda D, Kranda K, Bojar M, Glosová L, Bäurle J, Kenney J, et al. Antibody formation against β- tubulin class III in response to brain trauma. Brain Res Bull 2006; 68: 213–6.

Sorokina EG, Semenova ZhB, Bazarnaya NA, Meshcheryakov SV, Reutov VP, Goryunova AV, et al. Autoantibodies to Glutamate Receptors and Products of Nitric Oxide Metabolism in Serum in Children in the Acute Phase of Craniocerebral Trauma. Neurosci Behav Physiol 2009; 39: 329–34.

Stahel PF, Morganti-Kossmann MC, Perez D, Redaelli C, Gloor B, Trentz O, et al. Intrathecal levels of complement-derived soluble membrane attack complex (sC5b-9) correlate with blood-brain barrier dysfunction in patients with traumatic brain injury. J Neurotrauma 2001; 18: 773–781.

Steyerberg EW, Mushkudiani N, Perel P, Butcher I, Lu J, McHugh GS, et al. Predicting Outcome after Traumatic Brain Injury: Development and International Validation of Prognostic Scores Based on Admission Characteristics [Internet]. PLoS Med 2008; 5[cited 2019 Jun 19] Available from: https://www.ncbi.nlm.nih.gov/pmc/articles/PMC2494563/

Steyerberg EW, Wiegers E, Sewalt C, Buki A, Citerio G, De Keyser V, et al. Case-mix, care pathways, and outcomes in patients with traumatic brain injury in CENTER-TBI: a European prospective, multicentre, longitudinal, cohort study. Lancet Neurol 2019; 18: 923–34.

Sullivan GM, Mierzwa AJ, Kijpaisalratana N, Tang *Haiying, Wang Y, Song S-K, et al. Oligodendrocyte Lineage and Subventricular Zone Response to Traumatic Axonal Injury in the Corpus Callosum. J Neuropathol Exp Neurol 2013; 72: 1106–25.

Teasdale GM, Pettigrew LE l., Wilson J t. L, Murray G, Jennett B. Analyzing Outcome of Treatment of Severe Head Injury: A Review and Update on Advancing the Use of the Glasgow Outcome Scale. J Neurotrauma 1998; 15: 587–97.

Thelin E, Al Nimer F, Frostell A, Zetterberg H, Blennow K, Nyström H, et al. A Serum Protein Biomarker Panel Improves Outcome Prediction in Human Traumatic Brain Injury. J Neurotrauma 2019; 36: 2850–62.

Thelin EP, Zeiler FA, Ercole A, Mondello S, Büki A, Bellander B-M, et al. Serial Sampling of Serum Protein Biomarkers for Monitoring Human Traumatic Brain Injury Dynamics: A Systematic Review [Internet]. Front Neurol 2017; 8[cited 2019 Aug 28] Available from: https://www.ncbi.nlm.nih.gov/pmc/articles/PMC5494601/

Thompson HJ, Marklund N, LeBold DG, Morales DM, Keck CA, Vinson M, et al. Tissue sparing and functional recovery following experimental traumatic brain injury is provided by treatment with an anti-myelin-associated glycoprotein antibody. Eur J Neurosci 2006; 24: 3063–72.

Till C, Colella B, Verwegen J, Green RE. Postrecovery Cognitive Decline in Adults With Traumatic Brain Injury. Arch Phys Med Rehabil 2008; 89: S25–34.

Tiriveedhi V, Gautam B, Sarma NJ, Askar M, Budev M, Aloush A, et al. Pre-transplant antibodies to Kα1 tubulin and Collagen-V in lung transplantation: Clinical Correlations. J Heart Lung Transplant Off Publ Int Soc Heart Transplant 2013; 32: 807–14.

Werner C, Engelhard K. Pathophysiology of traumatic brain injury. BJA Br J Anaesth 2007; 99: 4–9.

Whitnall L. Disability in young people and adults after head injury: 5-7 year follow up of a prospective cohort study. J Neurol Neurosurg Psychiatry 2006; 77: 640–5.

Winter M, Baksmeier C, Steckel J, Barman S, Malviya M, Harrer-Kuster M, et al. Dose-dependent inhibition of demyelination and microglia activation by IVIG. Ann Clin Transl Neurol 2016; 3: 828–43.

Yue JK, Yuh EL, Korley FK, Winkler EA, Sun X, Puffer RC, et al. Association between plasma GFAP concentrations and MRI abnormalities in patients with CT-negative traumatic brain injury in the TRACK-TBI cohort: a prospective multicentre study. Lancet Neurol 2019; 18: 953–61.

Zhang Z, Zoltewicz JS, Mondello S, Newsom KJ, Yang Z, Yang B, et al. Human Traumatic Brain Injury Induces Autoantibody Response against Glial Fibrillary Acidic Protein and Its Breakdown Products. PLoS ONE 2014; 9: e92698.

